# Design of an optimal combination therapy with broadly neutralizing antibodies to suppress HIV-1

**DOI:** 10.1101/2021.12.03.21267269

**Authors:** Colin LaMont, Jakub Otwinowski, Kanika Vanshylla, Henning Gruell, Florian Klein, Armita Nourmohammad

## Abstract

Broadly neutralizing antibodies (bNAbs) are promising targets for vaccination and therapy against HIV. Passive infusions of bNAbs have shown promise in clinical trials as a potential alternative for anti-retroviral therapy. A key challenge for the potential clinical application of bnAbs is the suppression of viral escape, which is more effectively achieved with a combination of bNAbs. However, identifying an optimal bNAb cocktail is combinatorially complex. Here, we propose a computational approach to predict the efficacy of a bNAb therapy trial based on the population genetics of HIV escape, which we parametrize using high-throughput HIV sequence data from a cohort of untreated bNAb-naive patients. By quantifying the mutational target size and the fitness cost of HIV-1 escape from bNAbs, we reliably predict the distribution of rebound times in three clinical trials. Importantly, we show that early rebounds are dominated by the pre-treatment standing variation of HIV-1 populations, rather than spontaneous mutations during treatment. Lastly, we show that a cocktail of three bNAbs is necessary to suppress the chances of viral escape below 1%, and we predict the optimal composition of such a bNAb cocktail. Our results offer a rational design for bNAb therapy against HIV-1, and more generally show how genetic data could be used to predict treatment outcomes and design new approaches to pathogenic control.

## I. INTRODUCTION

Recent discoveries of highly potent broadly neutralizing antibodies (bNAbs) provide new opportunities to successfully prevent, treat, and potentially cure infections from evolving viruses such as HIV-1 [1–12], influenza [13], and the Dengue virus [14, 15]. bNAbs target vulnerable regions of a virus, such as the CD4 binding site of HIV *env* protein, where escape mutations can be costly for the virus [1–3, 16–19]. As a result, eliciting bNAbs is the goal of a universal vaccine design against the otherwise rapidly evolving HIV-1. Apart from vaccination, bNAbs can also offer significant advances in therapy against both HIV and influenza [15, 18, 20, 21]. Specifically, augmenting current anti-retroviral therapy (ART) drugs with bNAbs may provide the next generation of HIV therapies [21, 22].

Recent studies have used bNAb therapies to curb infections by the Simian immunodeficiency virus (SHIV) in non-human primates [23–25], and HIV-1 infections in human clinical trials [7–9, 26]. Monotherapy trials with potent bNAbs, including 3BNC117 [7], VRC01 [26], and 10-1074 [8] indicate that administering bNAbs is safe and can suppress viral load in patients. Nonetheless, in each trial, escape mutants emerge resulting in a viral rebound after about 20 days past infusion of the bNAb. However, in trials that administered a combination of 10-1074 and 3BNC117, viral rebound was substantially suppressed [9, 23]. Combination therapy has been repeatedly used against many infectious agents, including current HIV ART cocktails and combination antibiotic treatments against Tuberculosis [27]. The principle be-hind combination therapy is clear: It is harder for a pathogen population to acquire resistance against multiple treatment targets simultaneously than to acquiring resistance against each target separately.

Prior work has focused on optimizing combination therapy of bNAbs for breadth and potency [28, 29] without considering viral dynamics. Theoretical approaches have been used to model the dynamics of viremia in patients following passive infusion of bNAbs [30–33]. However the predictive power of models relying on trial data is limited by the small number of individuals enrolled in these trials, and increasing the size of a trial may be impractical. One can view the outcome of these trials in the context of an evolutionary competition among susceptible and resistant strains with different probabilities to emerge and establish in an HIV population within a patient, due to their different differential fitness effects in the presence or absence of a bNAb. Studies of population genetics of HIV have found rapid intra-patient evolution and turnover of the virus [34, 35] and have indicated that the efficacy of drugs in anti-retroviral therapy can severely impact the mode of viral evolution and escape [36]. Despite the complex evolutionary dynamics of HIV-1 within patients due to individualized immune pressure [37], genetic linkage [35], recombination [35, 38], and epistasis between loci [39, 40], the genetic composition of a population can still provide valuable information about the evolutionary significance of specific mutations, especially in highly vulnerable regions of the virus. For example, analysis of genomic covariation in the Gag protein of HIV-1 has been successful in predicting fitness effect of mutations in relatively conserved regions of the virus, which could inform the design of rational T-cell therapies that target these vulnerable regions [41].

Here, we present a statistical inference framework that uses the high throughput longitudinal survey of genetic data collected from 11 ART-naive patients over about 10 years of infection [35] to characterize the evolutionary fate of escape mutations and to predict patient outcomes in recent mono- and combination therapy trials with 10-1074 and 3BNC117 bNAbs [7–9]. Using the accumulated intra-patient genetic variation from deep sequencing of HIV-1 populations in ART–naive patients [35], we can estimate the diversity and the fitness effects of mutations at sites mediating escape. These variables parametrize our individual-based model for viral dynamics to characterize the expected path for a potential escape of HIV-1 populations in response to bNAb therapies in patients enrolled in the clinical trials. Our analysis accurately predicts the distribution of viral rebound times in response to passive bNAb infusions [7–9], measuring the efficacy of these clinical trials.

Our prediction for the viral rebound time in response to a bNAb is done based on the inferred genetics parameters from the deep sequencing of HIV-1 populations in a separate cohort of ART-naive patients. Therefore, we use our approach to assess a broader panel of nine bNAbs, for which escape sites can be identified from prior deep mutational scanning experiments [42], to characterize the therapeutic efficacy of each of these bNAbs and to propose optimal combination therapies that can efficiently curb an HIV infection. Our results showcase how the wealth of genetic data can be leveraged to guide rational therapy approaches against HIV. Importantly, this approach is potentially applicable to therapy designs against other evolving pathogens, such as resistant bacteria or cancer.

## II. MODEL

### HIV response to therapy

After infusion of bNAbs in a patient, the antibodies bind and neutralize the susceptible strains of HIV. The neutralized subpopulation of HIV no longer infects T-cells, and the plasma RNA copy-number associated with this neutralized population decays. The dynamics of viremia in HIV patients off ART following a bNAb therapy with 3BNC117 [7], 10-1074 [8], and their combination [9] are shown in Figs. S1-S3. With competition of the neutralized strains removed, the resistant subpopulation grows until the viral load typically recovers to a level close to the pretreatment state (i.e., the carrying capacity); see Fig. 1A. The time it takes for the viral load to recover is the *rebound time*– a key quantity that characterizes treatment efficacy within a patient. Although the details of the viremia dynamics, especially at beginning and at the end of the therapy, may be complex [30–33], the rebound time can be approximately modeled using a logistic growth after bNAb infusion (*t >* 0),

**FIG. 1.**
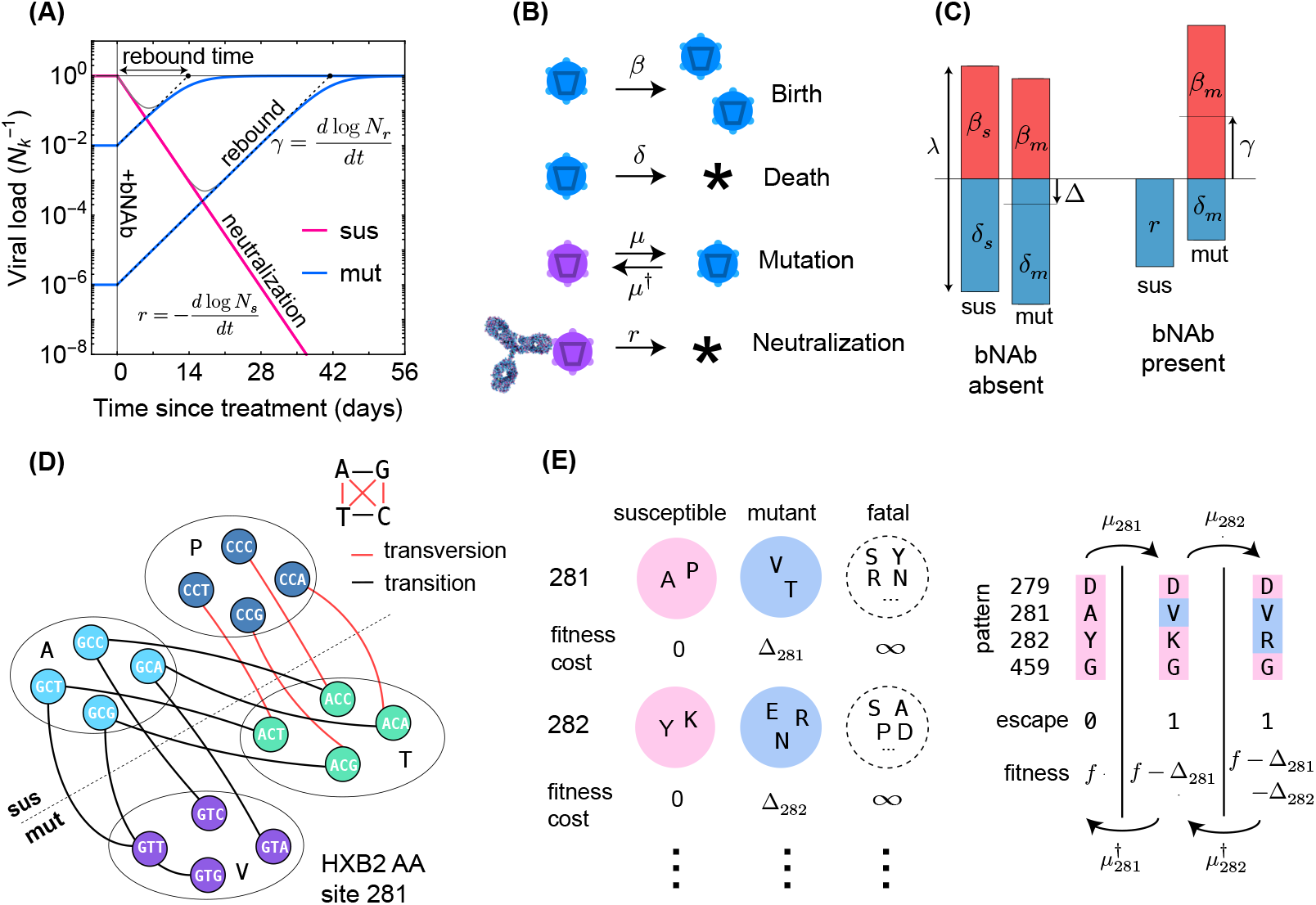
Schematics for the evolutionary dynamics of viral rebound. **(A)** The viral dynamics after the initiation of a treatment with bNAb infusion (*t* = 0) is determined by two competing processes. Susceptible strains (sus) undergo exponential decay (red line) with decay rate given by *r*, while the resistant mutants (mut) undergo logistic growth back up to the carrying capacity (*N*_*k*_) of the patient. In the deterministic limit (eq. 1), the rebound time is linearly related to the log-frequency of the mutant fraction. **(B)** The schematic shows the four stochastic processes of birth, death, mutation, and neutralization with their respective rates for susceptible (purple) and resistant (blue) variants. These processes define the evolution of a viral population. Note that both the susceptible and the resistant variants are subject to birth and death with their respective rates. **(C)** The birth and death rates can be visualized as a region of size *λ* = *β* + *δ* which is partitioned into birth and death events. In the absence of antibodies, the susceptible population has balanced birth and death rates, *β*_*s*_ = *δ*_*s*_, while the resistant population has a negative net birth rate equal to the fitness difference Δ = *δ*_*m*_*− β*_*m*_. After introduction of the antibody, the susceptible population decays at rate *r*, and without competition from the susceptible population, the resistant population grows at the free growth-rate *γ*. **(D)** Mutational target size is inferred *a priori* from the genotype-phenotype mapping, which can be visualized as a bipartite graph. The nodes correspond to codons, while the edges are the mutations which link one codon to another, weighted according to the respective mutation rates. The average edge weight from codons of susceptible variants to the escape mutants determines the rate of escape mutations *µ*. Mutations can be divided into two types: transitions (black) are within-class, and transversions (red) are out of class nucleotide changes. Transitions occur at about 8 times higher rate than transversions (Fig. 2). **(E)** A coarse grained fitness and mutation model for two of the escape sites (281 and 282) against antibody 3BNC117 are shown. Left: At each escape mediating site, amino acids fall into one of three groups: (i) susceptible (wild-type), (ii) escape mutant, and (iii) fatal. For an escape-class amino acid at site *i* the virus incurs a fitness cost Δ*i*, and these costs are additive across sites. Right: Mutations at a given site *i* occur with (independent) forward *µ*_*i*_ and backward 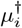 rates which govern the substitution events between amino acid classes.

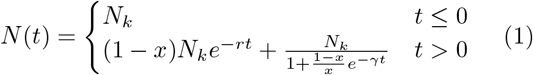

with the initial condition set for pre-treatment fraction of resistant subpopulation *x* = *N*_*r*_(0)*/*(*N*_*r*_(0) + *N*_*s*_(0)), where *N*_*r*_(0) and *N*_*s*_(0) denote the size of resistance and susceptible subpopulations at time *t* = 0, respectively. Here, *γ* is the growth rate of the resistant population, *r* is the neutralization rate impacting the susceptible sub-population, and *N*_*k*_ is the carrying capacity (Fig. 1A, Methods). In our analysis, we set *γ* = 1*/*3 days^*−*1^ or a doubling time of ∼ 2 days, which is the characteristic of HIV growth in patients [43]. We infer the neutralization rate *r* as a global parameter for each trial, since it depends on the neutralization efficacy of a bNAb at the concentration used in the trial. We will infer the patient-specific pre-treatment fraction of resistant subpopulation *x*, using a population genetics based approach based on which we characterize the mutational target size and selection cost of escape in the absence of a bNAb (see below). The resulting viremia fits in Figs. S1-S3 specify the rebound time *T* in each patient, which in this simple model, is given by *T* = −*γ*^*−*1^ log *x* (Methods).

The rebound time following passive infusion of 3BNC117 [7] and 10-1077 [8] bNAbs range from 1 to 4 weeks, with a small fraction of patients exceeding the monitoring time window in the studies (late rebounds past 56 days); see Figs. S1-S3. The distribution of rebound times summarizes the escape response of the virus to a therapy and directly relates to the distribution for the pre-treatment fraction of resistant variants *P* (*x*) across patients *P* (*T*) ∼ *x*^*−*1^*P* (*x*).

### Stochastic evolutionary dynamics of HIV subject to bNAb therapy

The fate of an HIV population subject to bNAb therapy depends on the composition of the pre-treatment population with resistant and susceptible variants, and the establishment of resistant variants following the treatment. To capture these effects, we construct an individual-based stochastic model for viral rebound (Fig. 1B). We specify a coarse-grained phenotypic model, where a viral strain of type *a* is defined by a binary state vector 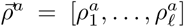, with *l* entries for potentially escape-mediating epitope sites; the binary entry of the state vector at the epitope site *i* represents the presence 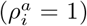 or absence 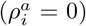 of a escape mediating mutation against a specified bNAb at this site of variant *a*. We assume that a variant is resistant to a given antibody if at least one of the entries of its corresponding state vector is non-zero.

At each generation, a phenotypic variant *a* can undergo one of three processes: birth, death and mutation to another type *b*, with rates *β*_*a*_, *δ*_*a*_, and *µ*_*a→b*_, respectively (Fig. 1B). The net growth rate of variant *a* is its birth rate minus the death rate, *γ*_*a*_ = *β*_*a*_ −*δ*_*a*_ (Fig. 1C). The total rate of events (birth and death) *λ* = *β*_*i*_ + *δ*_*i*_ modulates the amount of stochasticity in this birth-death process (Methods), which we assume to be constant across phenotypic variants. The continuous limit for this birth-death process results in a stochastic evolutionary dynamics for the sub-population of size *N*_*a*_,

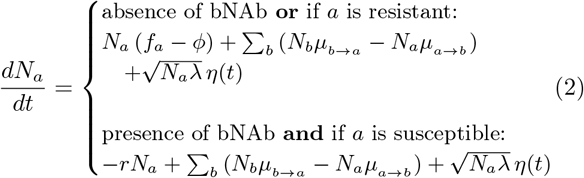

where *η*(*t*) is a Gaussian random variable with mean ⟨*η*(*t*) ⟩= 0 and correlation ⟨*η*(*t*)*η*(*t′*) ⟩= *δ*(*t t′*) (Methods). Here, *f*_*a*_ denotes the intrinsic fitness of variant *a* and its net growth rate *γ* is mediated by a competi-tive pressure 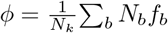 with the rest of the population constrained by the carrying capacity *N*_*k*_, such that *γ*_*a*_ = *f*_*a*_ *−ϕ*. In the presence of a bNAb, birth is effectively halted for susceptible variants and their death rate is set by the neutralization rate of the antibody, resulting in a net growth rate, *γ*_sus._ = *−r*. At the carrying capacity, the com<u>p</u>etitive pressure is the mean population fitness 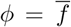, making the net growth rate of the whole population zero (Fig. 1C). When susceptible variants are neutralized by a bNAb, the competitive pressure *ϕ* drops, and as a result, the resistant variants can rebound to carrying capacity, at growth rates near their intrinsic fitness.

To connect the birth-death model (eq. 2) with data, we should relate the simulation parameters of a birthdeath process to molecular observables. We have already made a connection between the birth and death rates of a variant and its intrinsic fitness in eq. 2. In addition, the neutral diversity *θ* of a population at steady-state can be expressed as *θ* = 2*N*_*k*_*µ/λ*, where *µ* ≈10^−5^*/* day is the per-nucleotide mutation rate, which we infer from intra-patient longitudinal HIV sequence data [35] (Methods). For consistency, we set the total rate of events *λ* to be at least as large as the fastest process in the dynamics, which in this case is the growth rate of susceptible viruses *γ* ≈ (3 days)^*−*1^, we choose *λ* = (0.5 days)^*−*1^ (Methods). Therefore, the key parameters of the birth-death model, i.e., *β, δ*, and *N*_*k*_ can be expressed in terms of the intrinsic fitness of the variants *f*_*a*_ and the neutral diversity *θ*, which we will infer from data.

## III. RESULTS

### Population genetics of HIV escape from bNAbs

HIV escape from different bNAbs has been a subject of interest for vaccine and therapy design, and a number of escape variants against different bNAbs have been identified in clinical trials or in infected individuals [7–9, 44, 45]. This *in-vivo* data is often complemented with information from co-crystallized structures of bNAbs with the HIV envelope protein [46], and *in-vitro* deep mutational scanning (DMS) experiments, in which the relative change in the growth rate of tens of thousands of viral mutants are measured in the presence of different bNAbs [42, 47, 48]. We identify escape mutations against each of the bNAbs in this study by using information from clinal trials, the characterized binding sites, and the DMS assays (Methods); the list of escape mutations against each bNAb is given in Table S1.

The rise and establishment of an escape variant against a specific bNAb depend on three key factors, (i) neutral genetic diversity of the viral population, (ii) the mutational target size for escape from the bNAb and (iii) the intrinsic fitness associated with such mutations. Although viremia traces in clinical trials can be used to model the escape dynamics [30–33], they do not offer a comprehensive statistical description for HIV escape as they are limited by the number of enrolled individuals. Alternatively, mutation and fitness characteristics of such escape-mediating variants can be inferred from a broader cohort of untreated and bNAb-naive patients. We will infer these quantities from the large amount of high-throughput HIV sequence data from ref. [35] (see Fig. 2A, B for details) and use them to parameterize the birth-death model (Fig. 1B).

**FIG. 2.**
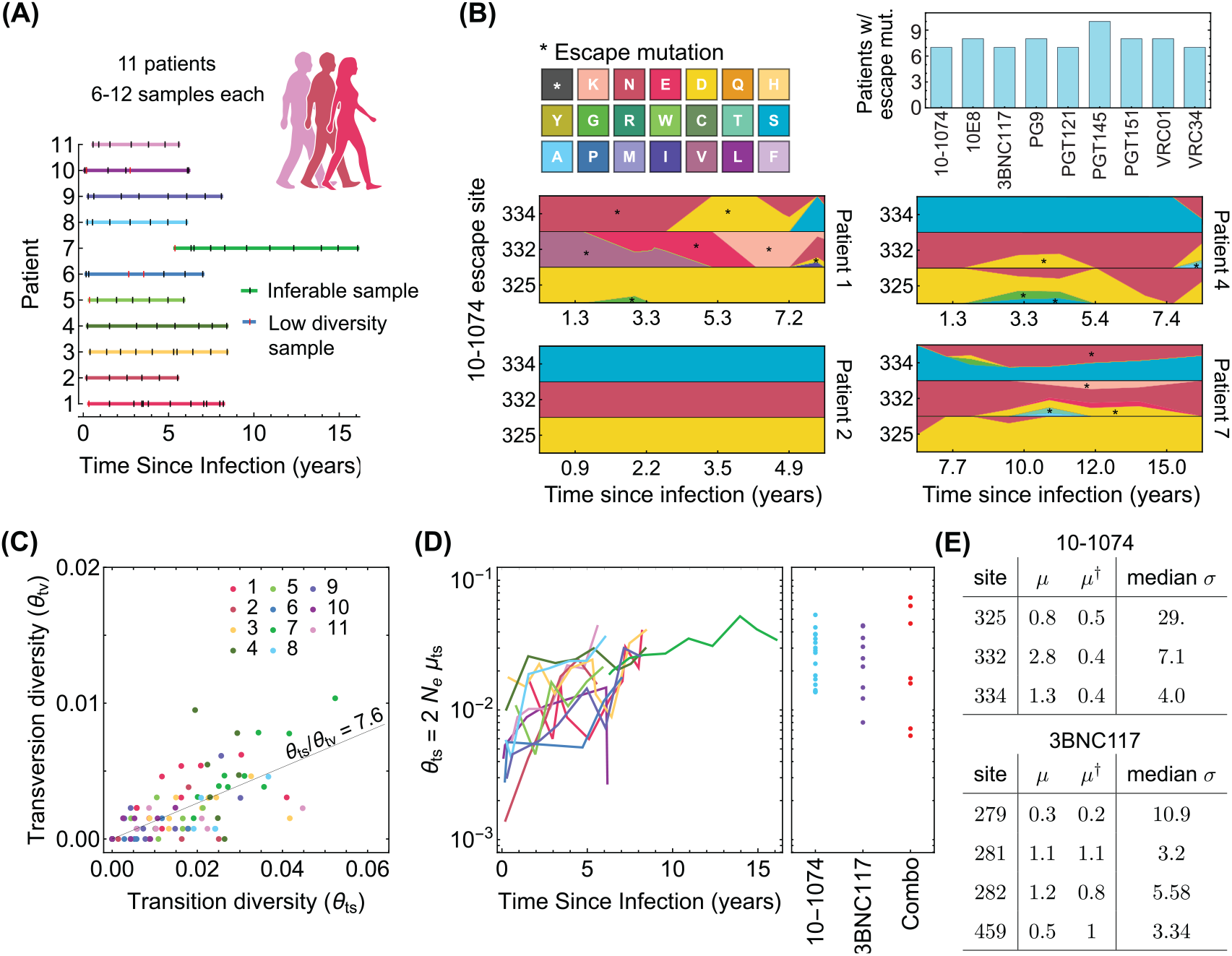
Statistics of the genetic data from bNAb-naive HIV patients. **(A)** Statistics of the high throughput longitudinal data collected from HIV populations in 11 ART-naive patients from ref. [35], is shown. Some of the have low diversity (vertical red lines) and were not usable for our study. Usable samples (vertical black lines) amount to 4-10 samples per patient, collected over 5 -10 years of infection. **(B)** Lower panels show the relative frequencies (cube-root transformed for legibility) of different amino acids in four patients at the 3 escape sites against the 10-1074 bNAb, estimated from the polymorphism data at the nucleotide level in each patient over time. Despite 10-1074 being a broadly neutralizing antibody, mutations associated with escape (indicated by a *∗*) are commonly observed in untreated patients. The upper right panel shows the number of individuals (out of the cohort of 11 bNAb-naive HIV patients) that carry mutations associated with escape against the indicated bNAbs. **(C)** The nucleotide diversity associated with transversion *θ*_tv_ = 2*N*_*e*_*µ*_tv_ is shown against the transition diversity *θ*_ts_ = 2*N*_*e*_*µ*_ts_ for all patients (colors) and all time points. The covariance of these two diversity measurements yields an estimate for the transition/transversion ratio *θ*_ts_*/θ*_tv_ = 7.6. **(D)** Left: The transition diversity is shown to grow as a function of time since infection in all the 11 patients (colors according to (A)). Right: The neutral diversity of viral populations in patients (points) from the three different clinical trials [7–9] analyzed in this study resemble the larger diversities of long-established viral populations in untreated patients. **(E)** The inferred forward and backward mutation rates (*µ, µ*^*†*^), relative to the transition rate, and the median selection strength *σ* at each escape site against the two bNAbs (10-1074, and 3BNC117) from the trial data used in this study are shown. Compared to the 10-1074 bNAb, escape from the 3BNC117 bNAb appears to be less costly, and is associated with a smaller mutational target.

### Diversity of the viral population

The neutral genetic diversity *θ* = 2*N*_*e*_*µ/λ* (i.e., the number of segregating alleles) determines the chance to observe a rare (e.g. resistant) mutation in a patient prior to treatment, and it modulates the strength of selection in an HIV population to escape a bNAb; here *N*_*e*_ is the effective population size, *µ* is the per-nucleotide mutation rate, and *λ* is the total number of events per virus in the birth-death process which determines the noise amplitude (Methods). We use synonymous changes as a proxy for diversity associated with the neutral variation in an HIV population at a given time point within a patient. By developing a maximum-likelihood approach based on the multiplicities of different synonymous variants, we can accurately infer the neutral diversity of a population from the large survey of synonymous sites in the HIV genome (Methods and Fig. S4). Importantly, we infer the neutral diversity of transition *θ*_ts_ and transversion *θ*_tv_ mutations sep-arately, and consistent with previous work [36, 49], find that transitions occur with a rate of about 8 times larger than transversions (Fig. 2C, Fig. S4B-E).

Our inference indicates that the neutral diversity grows over the course of an infection in untreated HIV patients from ref. [35] (Fig. 2D). The patients enrolled in the three bNAb trials [7–9] show a broad range of neutral diversity prior to bNAb therapy (Fig. 2D). In addition to the circulating viruses in a patient’s sera, the viral reservoir, which consists of replication-competent HIV in latently infected cells or un-sampled tissue, can also contribute to a bNAb escape in a patient. Evidence for the effect of reservoir is directly visible in trials as the failure of pre-trial sequencing to exclude patients who do not harbor escape variants. We model the effect of the reservoir as augmenting the neutral diversity by a constant multiplicative factor *r*_resv._, so that patients with more diverse sera, representing usually longer infections, are also expected to have correspondingly more diverse reservoir populations. By fitting the observed rebound data, we infer the reservoir factor *r*_resv._ ≃ 2.07 (Methods, Fig. S4). We use the augmented genetic diversity of HIV prior to the bNAb therapy in each trial to generate the rebound time and the probability of HIV escape in patients.

### Mutational target size for escape

We define the mutational target size for escape from a bNAb as the number of trajectories that connect the susceptible codon to codons associated with escape variants, weighed by their probability of occurrences (Methods). The connecting paths with only single nucleotide transitions or transversions dominate the escape and can be represented as connected graphs shown in Fig. 1D. To characterize the target size of escape for each bNAb, we determine the forward mutation rate *µ*≡ *µ*_sus.*→*res._ from the susceptible codons to the resistant (escape) codons, and the reverse mutation rate *µ*^*†*^ ≡ *µ*_sus._ ←_res._ back to the susceptible variant (Fig. 1D, Methods). The mutational target sizes vary across bNAbs, with HIV escape being most restricted from 10E8 (*µ/µ*_ts_ = 1.8) and most accessible in the presence of 10-1074 (*µ/µ*_ts_ = 4.9); see Fig. 4C and Table S1 for the list of mutational target size for escape against all bNAbs in this study.

### Fitness effect of escape mutations

Since bNAbs target highly conserved regions of the virus, we expect HIV escape mutations to be intrinsically deleterious for the virus [33, 41], and incur a fitness cost relative to pre-treatment baseline *f*_0_. We assume that fitness cost associated with escape mutations are additive and background-independent so the fitness of a variant *a* in the absence of bNAb follows, 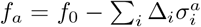, where Δ_*i*_ is the cost associated with the presence of a escape mutation at site *i* (i.e., for 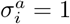); see Fig. 1D.

Interestingly, we observe the escape variants against different bNAbs to be circulating in the HIV populations from the cohort of ART- and bNAb-naive patients [35] (Fig. 2B). We use this data [35] and extract the multiplicity of susceptible and escape variants in HIV populations at each sampled time point from a given patient. We use a single locus approximation under strong selection to represent the stationary distribution of the underlying frequency of escape alleles *x* in each patient from ref. [35], 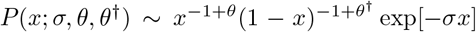, given the (scaled) fitness difference between the susceptible and the escape variants *σ* = 2*N*_*e*_(*f*_sus_ − *f*_mut_); see Methods.

Based on the statistics of escape and susceptible variants in all patients, we define a likelihood function that determines a Bayesian posterior for selection *σ* associated with escape at each site (Methods). We found that it is statistically more robust to infer the strength of selection relative to a reference diversity measure *σ/θ*_ts_ = (*f*_sus._ − *f*_res._)*/µ*_ts_, for which we choose the transition rate (Methods). This approach generates unbiased selection estimates in simulations and is robust to effects of linkage and recombination (Methods and Figs. S5, S6). The inferred values of the scaled fitness costs *σ/θ*_ts_ are shown for the escape-mediating sites of the trial bNAbs in Fig. 2E, and are reported in Table S1.

### Predicting the efficacy of bNAb therapy in clinical trials

Monotherapy trials with 10-1074 [8] and with 3BNC117 [7], and the combination therapy with both of these antibodies in [9] have shown variable outcomes. In some patients bNAb therapy did not suppress the viral load, whereas in others suppression was efficient and no rebound was observed up to 56 days after infusion (end of surveillance in these trials); see Fig. 3A for examples of patients with different rebound times, Figs. S1-S3 for the viremia traces in all patients, and Figs. 3B, C for the distributions and the summary statistics of the rebound times in patients in different trials.

**FIG. 3.**
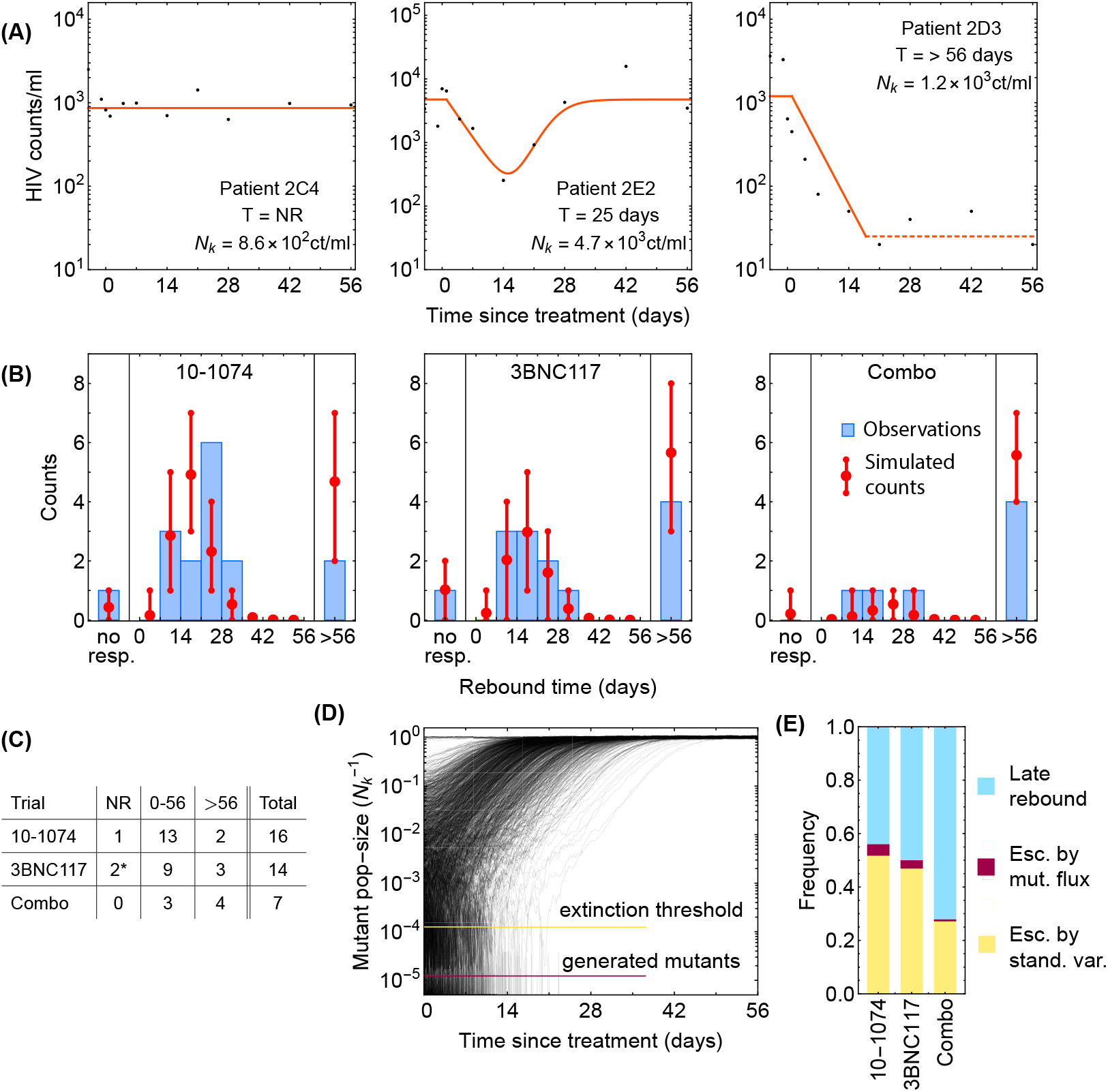
Statistics of viral rebound in clinical trials with bNAbs. **(A)** Panels show viremia of three patients from the 3BNC117 trial over time (black circles) and the fitted model of the viral decay and rebound processes from eq. 1 (orange line). The viral rebound time *T* and the fitted carrying capacity *N*_*k*_ is shown in each panel. Shown are examples of a non-responder (NR; left), a rebound occurring during the trial window (0 *< T <* 56 days; center), and a late rebound (*T >* 56 days; right). Viremia traces from all patients and all trials are shown in Figs. S1-S3. **(B)** We compare the distribution rebound times in patients from the three clinical trials with 10-1074 [8], 3BNC117 [7], and the combination of the two bNAbs [9] to the predictions from the simulations based on our evolutionary model (Fig. 1, and Methods). The error bars show the inter decile range (0.1-0.9 quantiles) generated by the simulations for the corresponding trial. **(C)** The summary table shows the number of patients for whom the infecting HIV population shows no response (NR), rebound during the trial window 0 *< T <* 56, and a late rebound (*T >* 56 days) in each trial. Note that 3 patients were excluded from the 3BNC117 trial (*) because of insufficient dosage leading to weak viral response: 1mg*/*kg compared to the 3*−* 30 mg*/*kg in the other treatment groups. **(D)** Plotted are 1, 200 trajectories of the mutant viral population simulated using our individual based model. Due to the individual birth-death events, fluctuations are larger when the population size is smaller. At a critical threshold, *x*_ext_, fluctuations are large enough to lead to almost certain extinction in the existing viral population. The critical threshold (yellow line) is an order of magnitude larger than the post-treatment spontaneously-generated mutant fraction (red line). **(E)** The predicted fraction of escape events associated with post-treatment spontaneous mutations (red) and the pre-treatment standing variation (yellow) are shown for the three trials. The fraction of events associated with late rebound is indicated in blue. Because the spontaneously-generated mutant fraction is smaller than the extinction threshold, these mutations contribute to less than 4% of escape events (red), and escape is likely primarily driven driven by standing variation (yellow), i.e., pre-existing escape variants.

**FIG. 4.**
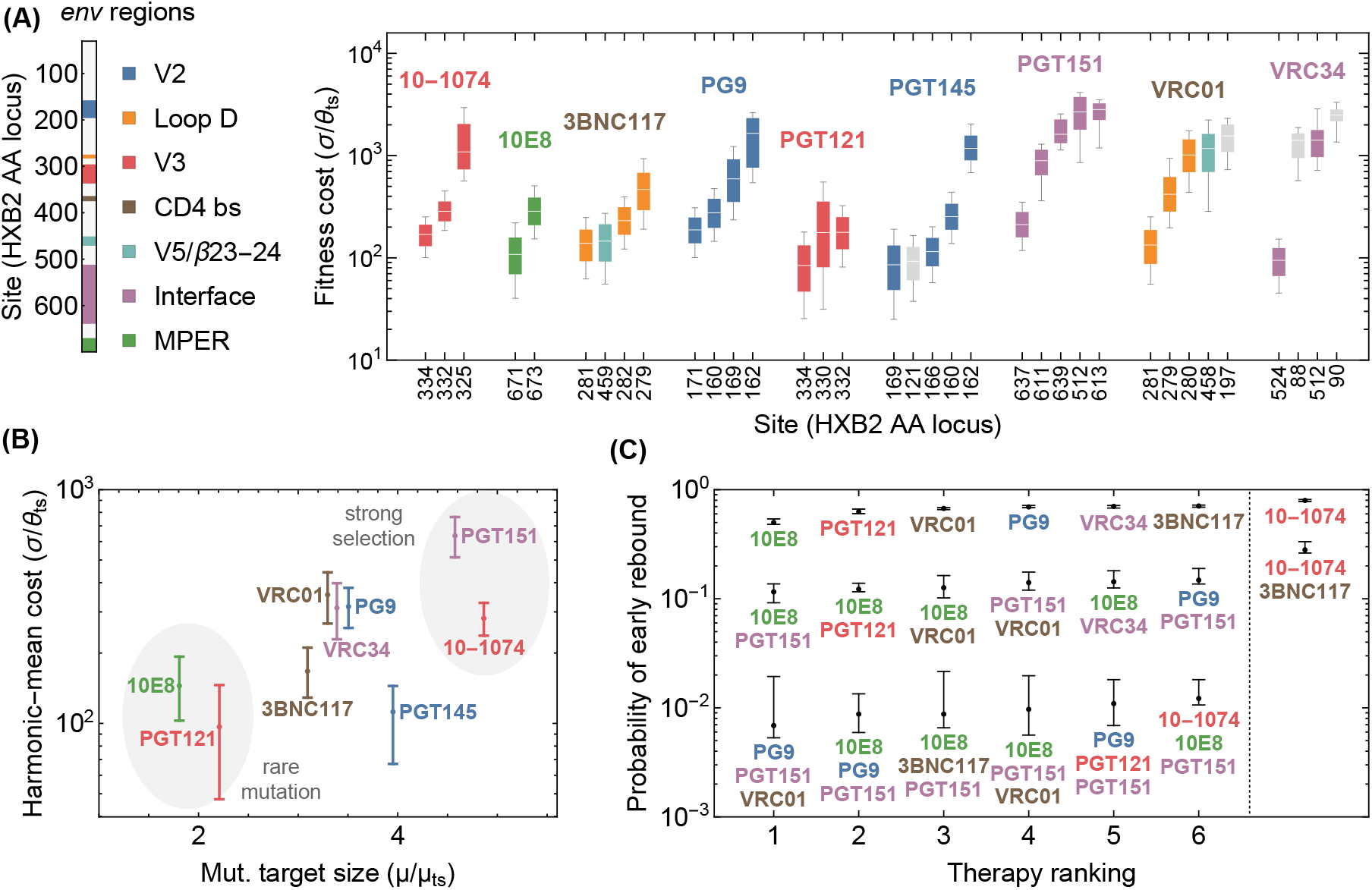
Statistics of viral escape for optimal combination therapy with bNAbs. **(A)** The posterior distribution for inferred selection strength on the escape-mediating sites associated with each of the 9 bNAbs in this study is shown (right); white line: median, box: 50% around the median, bar: 80% around median. Each escape site is color coded by its location on the *env* gene (left) and each antibody by its associated epitope location. **(B)** The harmonic mean of the selection strength *σ* associated with cost of escape (scaled by transition diversity *θ*_ts_) is shown against the mutational target size for each bNAb; error bars indicate 50% around the median. For antibodies to be broadly-neutralizing, it is sufficient that viral escape from them to be associated with a small mutational target size or a large fitness cost. The mutational target size is found to be weakly correlated with the average cost of escape from a given bNAb. We identify two distinct strategies for antibody breadth—selection limited and mutational-target-size limited escape pathways each highlighted in gray. **(C)** bNAb therapies with 1, 2, and 3 antibodies are ranked based on the predicted probability of early viral rebound, and in each case, six therapies with highest efficacies are shown; best ranked therapy is associated with the lowest probability of early rebound; indicate 50% around the median. Also for reference, the probability of early viral rebound two therapies from the trials in this study (10-1074 and 10-1074+3BNC117) are shown.

Although we infer a large intrinsic fitness cost for a virus to harbor an escape allele (Fig. 2E), these variants can emerge or already be present due to the large intra-patient diversity of HIV populations (Fig. 2C), or a larger mutational target size for these escape variants. Deep sequencing data in untreated (likely bNAb-naive) patients shows circulation of resistant variants against a panel of bNAbs in the majority of patients (Fig. 2B). Our goal is to predict the efficacy of a bNAb trial, using the fitness effect and the mutational target size for escape from a given bNAb, both of which we infer from the high-throughput HIV sequence data collected from bNAb-naive patients in ref. [35] (Figs. 2E, Table S1). In addition, we modulate these measures with the patient-specific neutral diversity *θ* inferred from whole genome sequencing of HIV populations in each patient prior to bNAb therapy (Fig. 2D). These quantities parametrize the birth-death process for viral escape in a bNAb therapy (Fig. 1A), which we use to characterize the distribution of rebound times in a given trial (Methods).

For both the the 3BNC117 and the 10-1074 trials [7, 8], we see an excellent agreement between our predictions of the rebound time distribution and data; see Fig. 3B, and Methods and Fig. S7 for statistical accuracy of this comparison. By assuming an additive fitness effect for escape from 10-1074 and 3BNC117, we also accurately predict the distribution of rebound times in the combination therapy [9] (Fig. 3B). The agreement of our results with data for combination therapy is consistent with the fact that the escape mediating sites from 10-1074 and 3BNC117 are spaced farther apart on the genome than 100bp, beyond which linkage disequilibrium diminishes due to frequent recombination in HIV [35]. Importantly, in all the trials, our evolutionary model accurately predicts the fraction of participants for whom we should expect a late viral rebound (more than 56 days passed bNAb infusion)—the quantity that determines the efficacy of a treatment.

Apart from the overall statistics of the rebound times, our stochastic model also enables us to characterize the relative contributions of the pre-treatment standing variation of the HIV population versus the spontaneous mutations emerging during a trial to viral escape from a given bNAb. Given the large population size of HIV and a high mutation rate (*µ* = 10^−5^nt per generation), spontaneous mutations generate a fraction *x*^(*µ*)^ of resistant variants during a trial, which we can express as,

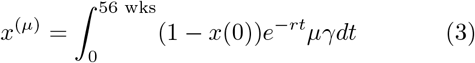

In the best case scenario, there are no resistant virions prior to treatment i.e., *x*(0) = 0. Since the neutralization rate *r* and the growth rate *γ* are comparable, this deterministic approach predicts that mutations can generate a resistant fraction of *x*^(*µ*)^ ≈10^−5^ during a trial. However, stochastic effects from random birth and death events play an important role in the fate and establishment of these resistant variants. The probability of extinction for a variant at frequency *x* can be approximated as 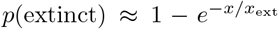 (Methods); here 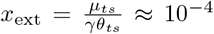 and a variant with fraction *x* that falls below this critical value is likely to go extinct (Fig. 3D). Since the total integrated mutational flux fraction during a trial is *x*^(*µ*)^ ∼10^−5^, mutational flux rarely decides the outcome of patient treatment. Indeed we infer that spontaneous mutations contribute to less than 4% of escape events in all the three trials and escape is primarily attributed to the standing variation prior from the serum or the reservoirs prior to therapy (Fig. 3E). A similar conclusion was previously drawn based on a mechanistic model of escape in VRC01 therapy trials [32].

## IV. DEVISING OPTIMAL BNAB THERAPY COCKTAILS

Clinical trials with bNAbs have been instrumental in demonstrating the potential role of bNAbs as therapy agents and in measuring the efficacy of each bNAb to suppress HIV. Still, these clinical trials can only test a small fraction of the potential therapies that can be devised. It is therefore important that trials test therapies that have been optimized based on surrogate estimates of treatment efficacy. The accuracy of our predictions for the rebound time of a HIV population subject to bNAb therapy suggests a promising approach to the rational design of therapies based on genetic data of HIV populations collected from bNAb-naive patients.

Here, we use genetic data to infer the efficacy of therapies with bNAbs, for which clinal trials are not yet performed. To do so, we first need to identify the routes of HIV escape from these bNAbs. We use deep mutational scanning data on HIV subject to 9 different bNAbs from ref. [42] together with information from literature to identify the escape mediating variants from each of these bNAbs (Methods and Table S1). We then determine the mutational target size and the fitness effect of these escape variants using high-throughput sequences of HIV in bNAb-naive patients from ref. [35]; these inferred values are reported in Fig. 4A, B and Table S1. Using the inferred fitness and mutational parameters and by setting the pre-treatment neutral diversity *θ* to be comparable to that of the patients in the previous three trials with 10-1074 and 3BNC117 (Fig. 2C), we simulate treatment outcomes for these 9 bNAbs and their 2-fold and 3-fold combinations.

Interestingly, we infer that escape from mono-therapies is almost certain and a combination of at least 3 antibodies is necessary to limit the probability of early rebound to below 1% (Fig. 4C). When considering all nine antibodies together, interesting patterns emerge. We find that the mutational target size and the fitness cost of escape, estimated as the harmonic-mean selection cost of individual sites, obey a roughly linear relationship (Fig. 4B). As all these bNAbs have similar overall breadth (i.e., they neutralize over 70% of panel strains), this result suggests that for an antibody to be broad, its escape mediating variants should either be rare (i.e, small mutational target size) or intrinsically costly (i.e., incurring a high fitness cost), but it is not necessary to satisfy both of these requirements. For instance, we find that 10E8 has a relatively weak selection but has a small escape target size, while PGT151 has a larger escape target size but makes up for it by having mutants with unavoidably high fitness cost.

The fitness-limited versus the mutation-limited strategies have different implications for the design of combination cocktails. The small mutational target size of 10E8 makes it the best candidate antibody for mono-therapy among the antibodies we consider because because the escape variants against this antibody are less likely to circulate in a patient’s serum prior to treatment. However, in combination, 10E8 appears less often in top ranked therapies than PGT151. PGT151 is unremarkable on its own because of a relatively large target size, but the high cost of escape makes it especially promising in combination therapies. Overall, fitness-limited bNAbs like PGT151 are more effective against high diversity viral populations, while mutation-limited bNAbs such as 10E8 are more effective against low diversity viral populations. Indeed, the best ranked therapy, namely the combination of PG9, PGT151, and VRC01, combines antibodies that target different regions of the virus and also have both types of fitness- and mutation-limited strategies for coverage against the full variability of viral diversities found in pre-treatment individuals (Fig. 4C) participating in the clinical trials.

## V. DISCUSSION

HIV therapy with passive bNAb infusion has become a promising alternative to anti-retroviral drugs for suppressing and preventing the disease in patients without a need for daily administration. The current obstacle is the frequent escape of the virus seen in mono- and even combination bNAb therapy trials [7–9, 26]. The key is to identify bNAb cocktails that can target multiple vulnerable regions on the virus in order to reduce the likelihood for the rise of resistant variants with escape-mediating mutations in all of these regions. Identifying an optimal bNAb cocktail can be a combinatorially difficult problem, and designing patient trials for all the potential combinations is a costly pursuit.

Here, we have proposed a computational approach to predict the efficacy of a bNAb therapy trial based on population genetics of HIV escape, which we parametrize using high-throughput HIV sequence data collected from a separate cohort of bNAb-naive patients [35]. Specifically, we infer the mutational target size for escape and the fitness cost associated with escape-mediating mutations in the absence of a given bNAb. These quantities together with the neutral diversity of HIV within a patient parametrize our stochastic model for HIV dynamics subject to bNAb infusion, based on which we can accurately predict the distribution of rebound times for HIV in therapy trials with 10-1074, 3BNC117 and their combination. Consistent with previous work on VRC01 [32], we found that viral rebounds in bNAb trials are primarily mediated by the escape variants present either in the patients’ sera or their latent reservoirs prior to treatment, and that the escape is not likely to be driven by the emergence of spontaneous mutations that establish during the therapy.

One key measure of success for a bNAb trial is the suppression of early viral rebound. Our model can accurately predict the rebound times of HIV subject to three distinct therapies [7–9], based on the fitness and the mutational characteristics of escape variants inferred from high-throughput HIV sequence data. This approach enables us to characterize routes of HIV escape from other bNAbs, for which therapy trials are not available, and to design optimal therapies. We used deep mutational scanning data [42] to identify escape-mediating variants against 9 different bNAbs for HIV. Our genetic analysis shows that bNAbs gain breadth and limit viral escape either due to their small mutational target size for escape or because of the large intrinsic fitness cost incurred by escape mutations. bNAbs with mutation-limited strategy are more effective at preventing escape in patients with low viral genetic diversity, while bNAbs selection-limited strategy with more effective at high viral diversity. To suppress the chance of viral rebound to below 1%, we show that a combo-therapy with 3 bNAbs with a mixture of mutation- and selection-limited strategies that target different regions of the viral envelope is necessary. Such combination can counter the full variation of viral diversity observed in patients. We found that PG9, PG151, and VRC01, which respectively target V2 loop, Interface, and CD4 binding site of HIV envelope, form an optimal combination for a 3-bNAb therapy to limit HIV-1 escape in patients infected with clade B of the virus.

We rest our analysis primarily on the predictive power of the observed variant frequencies in the untreated patients. Our model weighs these frequencies with respect to the viral diversity in a mathematically and biologically consistent way. However, we ignore the dynamics of antibody concentration and IC50 neutralization during treatments, the details of T-cell dynamics during infection [50], and also the evolutionary features of the genetic data, such as epistasis between loci [39, 40], genetic linkage [35], and codon usage bias [51]. The statistical fidelity of our model to the observed variant frequencies implies that many of the mechanistic details are at best secondary to the coarse-grained features of viral escape, and specifically the distribution of the rebound times. Nonetheless, our approach falls short of predicting the detailed characteristics of viremia traces in patients, especially at very short or very long times, during which the dynamics of T-cell response or the decay of bNAbs could play a role [30–32].

Our predictions are limited by our ability to identify the escape variants for each bNAb, either based on trial data, patient surveillance, or *in-vitro* assays such as DMS experiments. DMS data for viral escape [42, 47, 48] are generally of high quality but consider only a single HIV genetic background (e.g. in [47] this background is BF520.W14M.C2), whereas clinical data will have diverse baseline viruses between and within individuals. In addition, DMS data can be noisy for variants that grow very poorly in the absence of a bNAb, since growth without antibodies is the first stage of these experiments, followed by growth in the presence of bNAbs. This is likely to be the reason that the DMS experiments fail to capture a clear escape signal against bNAbs such as VRC01 or 3BNC117 that target the CD4 binding site of HIV, where mutations can be extremely deleterious in the absence of the bNAb. As such, we continue to need more data and more powerful statistical techniques for calling escape variants, using *in-vivo* approaches. For example, passive infusion of bNAb in humanized mouse models [52], which can, at least partially, reproduce the natural diversity of a human infection would be valuable. These efforts would take us a step closer to rational design for bNAb therapy and a more model-guided clinical trials.

Our approach showcases that, when feasible, combining high-throughput genetic data with ecological and population genetics models can have surprisingly broad applicability, and their interpretability can shed light into the complex dynamics of these populations. Application of similar methods to therapy design to curb the escape of cancer tumors against immune- or chemo-therapy, the resistance in bacteria against antibiotics, or the escape of seasonal influenza against vaccination is a promising avenue for future work. However, we expect that more sophisticated methods for inferring fitness from evolutionary trajectories may be necessary to capture the dynamical response of these populations.

## Data Availability

All data produced are available online at https://github.com/StatPhysBio/HIVTreatmentOptimization

## ACKNOWLEDGEMENTS

This work has been supported by the NSF CAREER award (grant No: 2045054), DFG grant (SFB1310) for Predictability in Evolution, and the MPRG funding through the Max Planck Society.

## Supplementary Information

### Data and code accessibility

The code for the algorithms used in this work and the data are available on GitHub at https://github.com/StatPhysBio/HIVTreatmentOptimization and in the Julia package https://github.com/StatPhysBio/EscapeSimulator.

## 1 Description of molecular data

### Data from bNAb trials

In this study we considered three clinical trials for passive therapy with bNAbs:

- Monoclonal therapy with 3BNC117 bNAb [1] with 16 patients enrolled, 13 of whom were off anti-retroviral therapy (ART).
- Monoclonal therapy with 10-1074 bNAb [2], with 19 patients enrolled, 16 of whom were off ART.
- Combination therapy with 10-1074 + 3BNC117 bNAb [3], with 7 patients off ART.

All sequenced patients across all trials were infected with distinct HIV-1 clade B viral strains. We limited our analyses to those patients not on ART at the time of treatment initiation. In these studies, the injected bNAb level falls off over time within patients and therefore, we only considered dynamics within an 8 week window since infusion. This assures that rebound is not confounded by a drop in bNAb below sensitive-strain neutralizing levels of 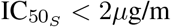.

We used single-genome sequence data of *env* collected from all patients in each trial to characterize the diversity of HIV population within each patient shown in Fig. 2 available from European Nucleotide Archive (Accession no: PRJEB9618). The patient sequence data for each trial is available through ref. [1] GeneBank PopSet: 1036347437, ref. [2] GenBank accession numbers KY323724.1 - KY324834.1, and ref. [3], GenBank accession numbers MH632763 - MH633255.

### Longitudinal HIV sequence data from untreated patients

Single nucleotide polymorphism (SNP) data was obtained from ref. [4] and aligned to the HXB2 reference using HIV-align tool [5]. The dataset includes 11 patients observed for 5-8 years of infection, with HIV sequence data sampled over 6-12 time points per patient (Fig. 2).

Patient 4 and patient 7 were excluded from the original analysis done in ref. [4] because of suspected superinfection and failure to amplify early samples, respectively. These patients were included in our analysis, since (i) super-infection poses no additional difficulties for our tree-free procedure, and (ii) only time points with measurable viral diversity entered into our selection likelihood, which automatically limits our analysis to samples with high quality sequences.

All patients were infected with clade B of HIV, except for patient 6 (clade C), and patient 1 (clade 01 AE). We assessed the robustness of our inference to exclusion of these patients from our analysis in Section 6. Overall, our inference was not strongly affected by this choice (Fig. S5), and therefore, we included these patients in our main analyses to enhance the statistics with larger data.

For our analysis we considered only data reported in single nucleotide polymorphism (SNP) counts. The number of raw SNP counts are the result of amplification and must be converted into estimates for the number of pre-amplification template fragments. Zanini et. al. reported that on average about 10^2^ templates of amplicon were associated with the fragments of the envelope (*env*) protein [4]. We converted raw SNP counts into templates by normalizing to 120 counts and rounding the resulting number to the nearest integer.

## 2 Identifying escape-mediating variants against bNAbs

The starting point for our analysis of HIV response to a given a bNAb is the description of the escape-mediating amino acids in the HIV *env* protein. We use a combination of methods to identify the escape variants for a given bNAb. First, we use deep mutational scanning (DMS) data of HIV-1 in the presence of a bNAb from ref. [6] to identify these mutations. These DMS experiments have created libraries of all single mutations from a given genomic background of HIV and tested the fitness of these variants (i.e., growth on T-cell culture) in the absence and presence of 9 different bNAbs, including the 10-1074 and 3BNC117 [6].

Escape variants in DMS data are identified as those which are strongly selected for only in the presence of a bNAb. Specifically, we identify escape variants as those which show 3-logs-change in their frequency in the presence versus absence of a bNAb. DMS data reflects *in-vitro* escape in cell culture. However, some of these variants may not be viable in vivo. To identify the reasonable candidates of escape in vivo, we limit our set to the variants that are also observed in the circulating viral strains of untreated HIV-1 patients from ref. [4]. It should be noted that since we use HIV sequence data from ref. [4] to infer selection on escape mediating variants in the absence of a bNAb, the candidates of escape that are not observed in the dataset [4] would be inferred to be strongly deleterious, and hence, unlikely to contribute to our predictions of viral rebound.

Our analysis of DMS data results in a set of escape mediating amino acids for 10-1074 that is consistent the escape variants that emerge in response to the bNAb trial [2] (Table S1). However, the DMS data is very noisy for bNAbs that target CD4 binding site of HIV, i.e., 3BNC117 and VRC01 [6]. One reason for this observation may be that the CD4 binding site is crucial for the entry of HIV to the host’s T-cells and mutations in this region are highly deleterious. As a results, only a small number of variants with mutations in this region can survive in the absence of a bNAb in a DMS experiment. Growth in the absence of a bNAb is the first step in the DMS experiments, which is then followed by exposure of the replicated variants to a bNAb. Therefore, a low multiplicity of variants in the absence of a bNAb could result in a noisy pattern of growth of the small subpopulation in the next stage of the experiment, in which growth is subject to a CD4-targeting bNAb.

For the CD4 binding site antibodies 3BNC117 and VRC01, we used additional data to call the escape variants. For 3BNC117 we used a combination of trial-patient sequences [1, 7], i.e. post-treatment enrichment, along with contact site information compiled in the crystallographic studies to narrow down candidate sites [8, 9]. For VRC01 we assumed a similar escape pattern to 3BNC117 but included sites known from other studies [10] and the clear DMS signal at HXB2 site 197. The sites we called were similar to those identified using humanized-mouse models of HIV infection [11], although more complex mutational patterns were seen in the soft-randomization scanning of [12]. Although the complete list of escape substitutions are unknown and background-dependent [12], the escape profiles which are most important are those that are most likely to be seen consistently in data and to be correctly identified. The list of substitutions are shown in Table S1.

## 3 Statistics and dynamics of viral rebound

### 3.1 Inference of growth parameters from dynamics of viremia

The concentration of viral RNA copies in blood serum is a delayed reflection of the total viral population size *N* (*t*), containing a resistant and susceptible subpopulations, with respective sizes *N*_*r*_(*t*) and *N*_*s*_(*t*). After infusion of bNAbs in a patient, the susceptible sub-population decays due to neutralization by bNAbs and the resistant sub-population grows and approaches the carrying capacity *N*_*k*_, with the dynamics,

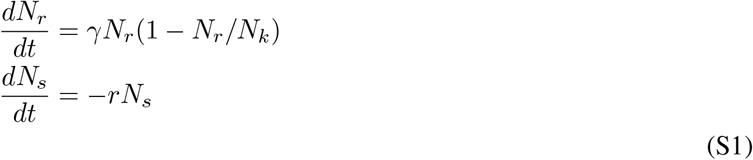

Here, *γ* is the growth rate of the resistant population, and *r* is the neutralization rate impacting the susceptible subpopulation. By setting the initial condition for fraction of resistant subpopulation prior to treatment (at time *t* = 0) *x* = *N*_*r*_(0)*/*(*N*_*r*_(0) + *N*_*s*_(0)), we can characterize the evolution of the total viremia in a patient. This dynamics is governed by the combined processes of neutralization by the infused bNAb and the viral rebound (Fig. 1A), which entails,

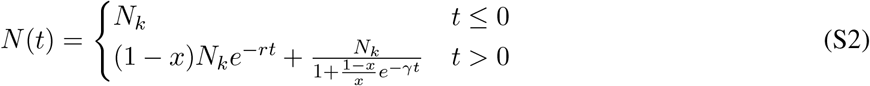

We use eq. S2 to define the evolution of blood concentration of viral RNA sequences which is observed indirectly via noisy viremia measurement data from refs. [1–3]. To connect the data with the simple model of viral dynamics in eq. S2, we fit the initial frequency of resistant mutants *x* for each patient separately, and fit a global estimate for the decay rate of susceptible variants *r* shared across all patients in a trial, using a joint maximum-likelihood procedure. In addition, we fix the growth rate *γ* to 1/3 days, corresponding to a doubling time of approximately 2 days [13]. Our analyses indicate that the initial viremia decline lagged treatment by about 1 day (Figs. S1-S3), consistent with previous findings [14]), and therefore, we included a 1-day lag between the fitted viremia response model and the treatment.

The number of viral RNA copies in a blood sample is subject to count fluctuations with respect to the true number of circulating virions in a given volume of the blood. We use a Poisson sampling model to define a likelihood for our model of viral population. The likelihood of observing *k* = *n*_*p*_(*t*) viral counts in a sample collected from patient *p* at time *t* is given by a Poisson distribution,

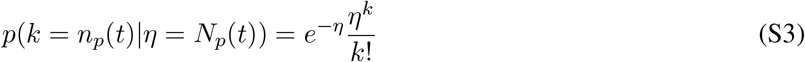

with rate parameter set by the model value of the viral multiplicity *N*_*p*_(*t*) (eq. S2). We use the Poisson likelihood in eq. S3 to characterize an error model to fit the parameters of the viral dynamics in eq. S2. However, since the mean and variance of the Poisson distribution are related, combining data with different mean values *N*_*p*_(*t*) at different times and from different patients can cause inconsistencies in evaluations of errors in our fits. To overcome this problem, we use a variance stabilizing transformation [15] and define a change in variable 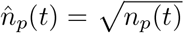. This transformed variable has a constant variance, a<u>n</u>d in the limit of large-sample size, it is Gaussian distributed with a mean and avariance given by, 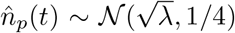. The constant variance of the transformed variable enables us to combine data from all patients and time points, irrespective of the sample’s viral loads, and fit the model parameters (*r, x*_*p*_) using (non-linear) least-squares fitting of the function

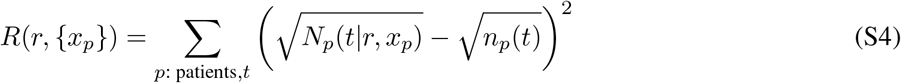

Here, *N*_*p*_(*t* | *r, x*_*p*_) is the model estimate of viremia in patient *p* at time *t* (eq. S2), given the pre-treatment fraction of resistant variants *x*_*p*_, and the decay rate *r*.

Note that the viremia measurements have a minimum sensitivity threshold of 20 RNA copies per ml. We treat the data points below the threshold of detection as missing data and if *n*_*p*_(*t*) is below the threshold of detection we impute *n*_*p*_(*t*) = min(20, *N*_*t*_).

The fitted viremia curves for patients enrolled in the three bNAb trials under consideration are shown in Figs. S1-S3, and the respective decay rates *r* for each experiment are,

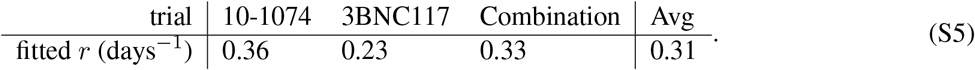

### 3.2 Individual-based model for viral population dynamics

To encode for different viral variants, we specify a coarse-grained phenotypic model, where a viral strain of type *a* is defined by a binary state vector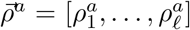, with *l* entries for potentially escape-mediating epitope sites; the binary entry of the state vector at the epitope site *i* represents the presence 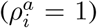 or absence 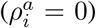 of a escape mediating mutation against a specified bNAb at site *i* of variant *a*. We assume that a variant is resistant to a given antibody if at least one of the entries of its corresponding state vector is non-zero.

We define an individual-based stochastic birth-death model [16] to capture the competitive dynamics of different HIV variants within a population. This dynamic model will allow us to predict the distribution of rebound times under any combination of antibodies.

We assume that a viral strain of type *a* can undergo one of three processes: birth, death and mutation to another type *b* with rates *β*_*a*_, *δ*_*a*_, and *µ*_*a→b*_, respectively:

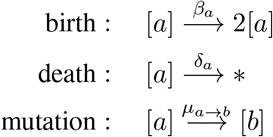

We specify an intrinsic fitness *f*_*a*_ for a given variant *a*, defined as the growth rate of the virus in the absence of neutralizing antibody or competition. Since bNAbs target highly vulnerable regions of the virus, we expect that HIV escape mutations to be intrinsically deleterious for the virus and to confer a fitness cost relative to the susceptible viral variants prior to the infusion of bNAbs. Assuming that fitness cost of escape is additive across sites and background-independent, we can express the fitness of a variant as, 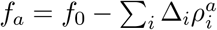, where Δ_*i*_ is the cost associated with the presence of an escape mutation at site *i* of variant *a* (i.e., for 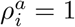).

We assume that growth is self-limiting via a competition for host T-cells. This competition enforces a carrying capacity, which sets the steady-state population size *N*_*k*_. Competition is mediated through a competitive pressure term 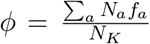 which attenuates the net growth rate *γ*_*a*_ so that *γ*_*a*_ = *f*_*a*_ − *ϕ*. At the carrying capacity, the competitive pressure equals the mean population fitness 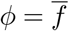, making the net growth rate of the population zero. The net growth rate of a variant *a* is given by its birth rate minus the death rate: *γ*_*a*_ = *β*_*a*_ *−δ*_*a*_. We assume that the total rate of events (i.e., the sum of birth and death events) is equal for all types, i.e., *λ* = *β*_*a*_ + *δ*_*a*_, ∀*a*. Assuming that *λ* is constant is to be agnostic about the mechanism of a fitness decrease, attributing fitness loss equally to (i) an increase in the death rate, and (ii) a decrease in the birth rate.

Because the absolute magnitude of *β* and *δ* asymptotically converge in the continuum limit for a surviving population, i.e., lim_*N→∞*_ *β/δ* = 1, it is impossible to distinguish between (i) and (ii) in the continuous limit.

Choosing constant *λ* simplifies both theoretical calculations and the simulation algorithm.

This leads to the following equations for the birth and the death rates:

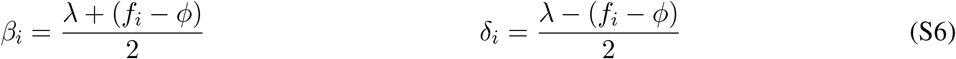

In the presence of an antibody, birth is effectively halted for susceptible variants, resulting in birth and death rate for a susceptible variant *s*,

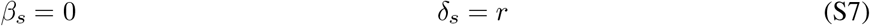

so that the susceptible phenotype decays at rate *r*.

We assume that mutations occur independently at each site,

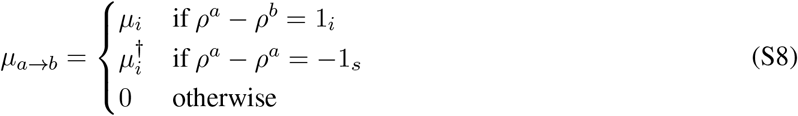

where 1_*s*_ is the vector which has only one non-zero entry at site *i*, and *µ*_*i*_ and 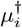 are the forward the backward mutation rates at site *i*, respectively.

We characterize the state of a population by vector ***n*** = (*n*_1_, … *n*_*M*_), where *n*_*a*_ is the number of type *a* variants within the population. The individual-based birth-death model introduced above specifies the stochastic dynamics of a population state over time. Using the concept of chemical reactions, suitable for Gillespie algorithm [16, 17], we can determine the propensity *a*_*r*_(***n***) for a given reaction *r* (i.e., birth, death, or mutation) in a population of state ***n***, which in turn determines the rate at which the reactions occur (eq. S6). We denote the resulting change in the state of a population due to reaction *r* by ***ν***_*r*_. Taken together, the impact of the reactions in the birth-death model can be summarized as,

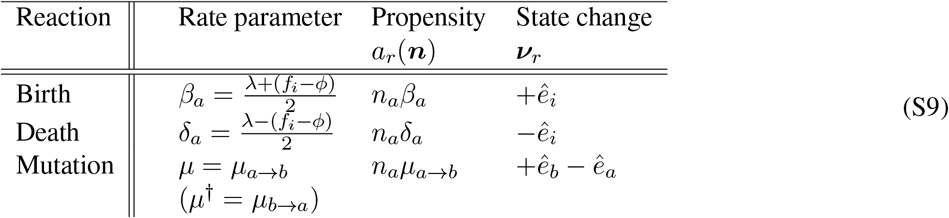

where 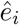 is a vector of size *M* equal to size of the population state vector, in which the *i*^*th*^ element equal to one and the rest are zero. For example, a mutation reaction *µ*(*a→ b*) destroys a variant *a* and creates a variant *b*, resulting in the following change in the state vector,

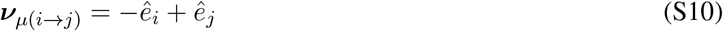

The reactions in eq. S9 specify a Master equation for the change in the probability of the population state *p*(***n***),

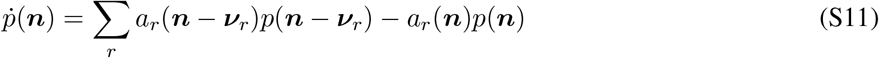

where 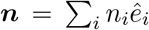 is the state vector. Using a Kramers-Moyal expansion [18], we arrive at a Fokker-Planck approximation for the change in the probability distribution of the population state *p*(***n***),

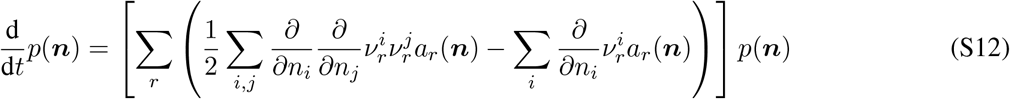

We can identify the drift (i.e., the deterministic force) and diffusion tensors of the Fokker-Planck operator:

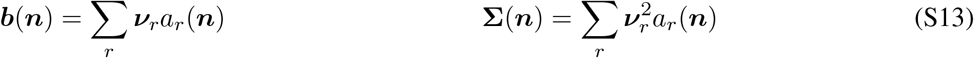

To better demonstrate the structure of this birth-death operator, consider a bi-allelic case (e.g. susceptible and resistant) with a 2-dimensional state vector, 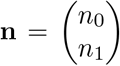. The drift and the diffusion tensors associated with this process follow,

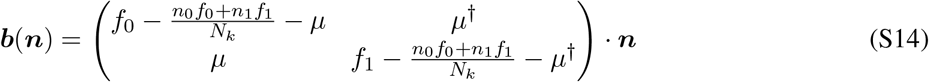

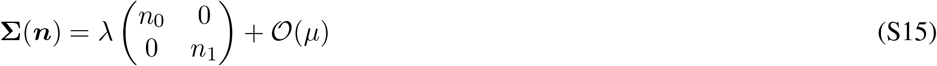

Note that in eq. S15 we neglect the stochasticity due to mutations since the magnitude of the associated noise is much smaller than the noise due to the birth and death (i.e., genetic drift).

Of special interest for our analysis is the steady-state density of frequencies, which we use to describe the initial state of the population (before treatment) and to infer selection intensity. In the steady state, the population is fluctuating around carrying capacity ∑ _*a*_ *n*_*a*_ ≈ *N*_*k*_ and we can represent the population state via allele frequencies *x*_*a*_ = *n*_*a*_*/N*_*k*_. In the simple case of a bi-allelic problem, the equilibrium allele frequency distribution *P*_eq_(*x*) follows the Wright-equilibrium distribution [19] with modified rates,

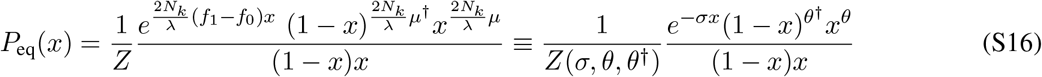

where *Z* is the normalization factor, *f*_1_ is the intrinsic fitness of the variant of interest, *f*_0_ is the fitness of the competing variant, *µ* and *µ*^*†*^ are the forward and backward mutation rates, *N*_*k*_ is the carrying capacity, and *λ* is the total rate of events in the birth-death process, which sets a characteristic time scale over which the impact of selection and mutations can be measured. In this case, we can define an “effective population size” that sets the effective size of a bottleneck and the natural time scale of evolution as *N*_*e*_ = *N*_*k*_*/λ*, and specify a scaled selection factor *σ* = *N*_*e*_*s* = *N*_*e*_(*f*_0_*− f*_1_), and scaled forward mutation and backward mutation rates (diversity) *θ* = 2*N*_*e*_*µ*, and *θ*^*†*^ = 2*N*_*e*_*µ*^*†*^. The normalization factor is given by,

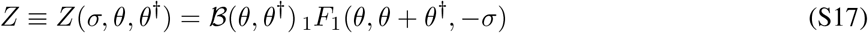

where _1_*F*_1_(·) denotes a Kummer confluent h ypergeometric function and 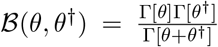 is the Euler beta function.

### 3.3 Extinction Probability

The logistic dynamics describing a patient’s viremia over time in eq. S2 is the deterministic approximation to the underlying birth-death process. However, the resistant population can also go extinct due stochastic effects, which in turn contribute to the probability of late rebound in a population. To capture this effect, we derive an approximate closed form expression for the probability of extinction.

Using the standard birth-death process generating function theory [20] the probability *P* (extinct |*n*_*i*_) that a population consisting of *n*_*i*_ resistant variants of type *i* go extinct can be expressed as,

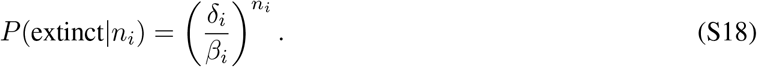

To characterize the probability of extinction for a population of size *N*_*k*_ with pre-treatment fraction of *i*^*th*^ resistant variants *x*_*i*_, we can convolve the extinction probability in eq. S18 with a Binomial probability density for sampling *n*_*i*_ resistant variants from *N*_*k*_ trials. Given that the pre-treatment fraction of resistant variants is small *x*_*i*_ *&$x226A;*1 and *N*_*k*_ is large, this Binomial distribution can be well approximated by a Poisson distribution, Poiss(*n*_*i*_; *N*_*k*_*x*_*i*_) with rate *N*_*k*_*x*_*i*_, resulting in an extinction probability,

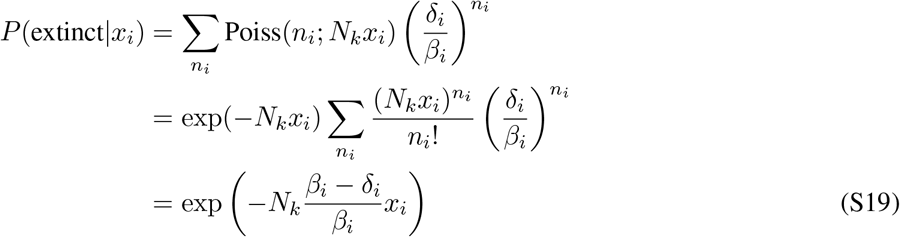

Using the expressions for the growth in the absence of competition, *β*_*i*_ − *δ*_*i*_ = *γ*_*i*_ = *f*_*i*_ (since *ϕ* = 0), and assuming that fitness is small relative to the total rate of birth and death events *f*_*i*_ *≪ λ*, we can use the approximation *β*_*i*_ = (*λ* + *f*_*i*_)*/*2 ≈ *λ/*2, to arrive at,

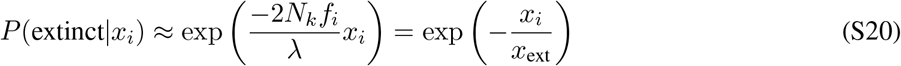

Where the characteristic escape threshold *x*_ext_ can be written in terms of concrete genetic observables,

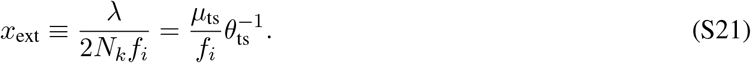

Fig. 3 shows that this threshold can well separate the fate of stochastic evolutionary trajectories, simulated with relevant parameters for intra-patient HIV evolution.

### 3.4 Numerical simulations of the birth-death process

To treat the full viral dynamics including mutations, and transient competition effects, we can exactly simulate the viral dynamics defined by our individual based model. Below are the key steps in this simulation.

#### Population initialization

At the starting point, we set the population size (i.e., the carrying capacity in the simulations) *N*_*k*_ as a free parameter chosen to be large enough to make discretization effects small. The population is then evolved through time using an exact stochastic sampling procedure (the Gillespie algorithm [17]). Simulating the outcome of this stochastic evolution generates the distribution of rebound times and the probability of late rebound—the key quantities related to treatment efficacy.

The input to our procedure is a list of antibodies for which we specify (i) the escape mediating sites for each antibody, and the (invariant) quantities describing (ii) the site-specific cost of escape 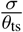, and (iii) the forward and backward mutation rates 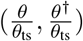. To simulate the trial outcome for each patient, we use the neutral population diversity *θ*_ts_ directly inferred from the patients (see Sec. 4.1). From this, we construct the list of *L* site parameters (concatenated across all antibodies) for selection and diversity: 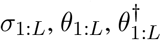.

We assume that at the start of the simulation, populations are in the steady state and that the potential escape sites are at linkage equilibrium. The approximate linkage equilibrium assumption is justified since the distance between these escape sites along the HIV genome is greater than the characteristic recombination length scale ≈ 100bp of the virus [4]. As a result, we draw an independent frequency *x*_*i*_ from the stationary distribution 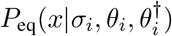 in eq. S16 to describe the state of a give site *i*, and use these frequencies to construct the initial viral genotypes *ρ*^*v*^ for each virus *v* in our initial population (Algorithm 1). In simulations, we show that this assumption does not bias our results even when *θ*_ts_ is fluctuating and recombination is absent (Section 6 and Fig. S6).

##### Algorithm 1

Population initialization

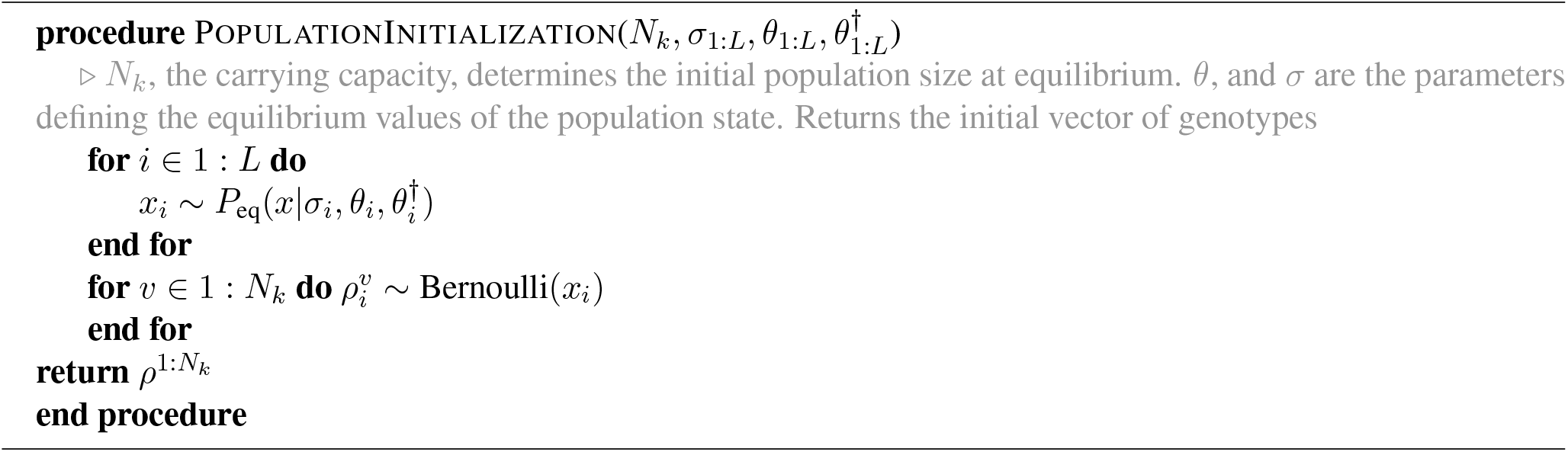

To sample from the stationary distribution itself, we define a novel Gibbs-sampling procedure [21] for generating the allele frequencies of the escape variants for the initial state of the population *x* ∼ *P*_eq_(*x* |*σ, θ, θ*^*†*^) (eq. S16). To characterize this procedure, we expand the exponential selection factor *e*^*σ*(1*−x*)^ in the original distribution, which results in,

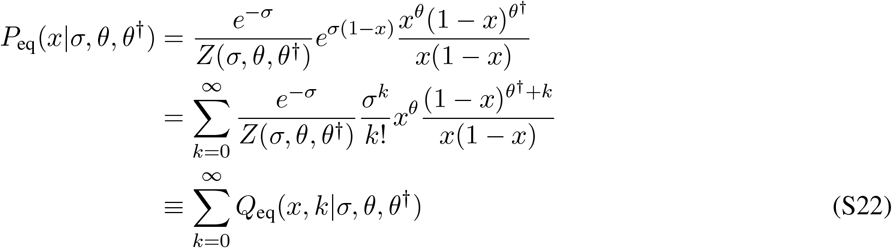

Here, *Q*_eq_(*x, k* |*σ, θ, θ*^*†*^) is a joint distribution over (*x, k*), and the desired distribution over the allele frequency *x* can be achieved by marginalizing the joint distribution over the discrete variable *k*. We can also express the conditional distributions for *x* and *k* as,

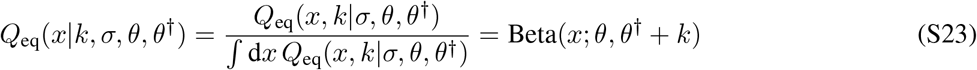

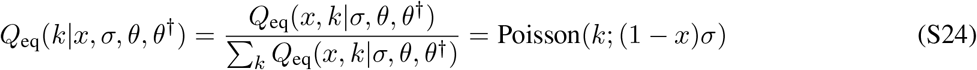

We use these conditional distributions to define a joint Gibbs sampler for *Q*_eq_. We summarize the resulting (*x, k*) ∼*Q*_eq_(*x, k σ, θ, θ*^*†*^) in the joint Gibbs sampler in Algorithm 2. This chain mixes extremely quickly and avoids calculation of the hypergeometric function for the normalization factor (eq. S24), which is computationally costly (Algorithm 2).

##### Algorithm 2

Gibbs Sampler for Allele Frequencies

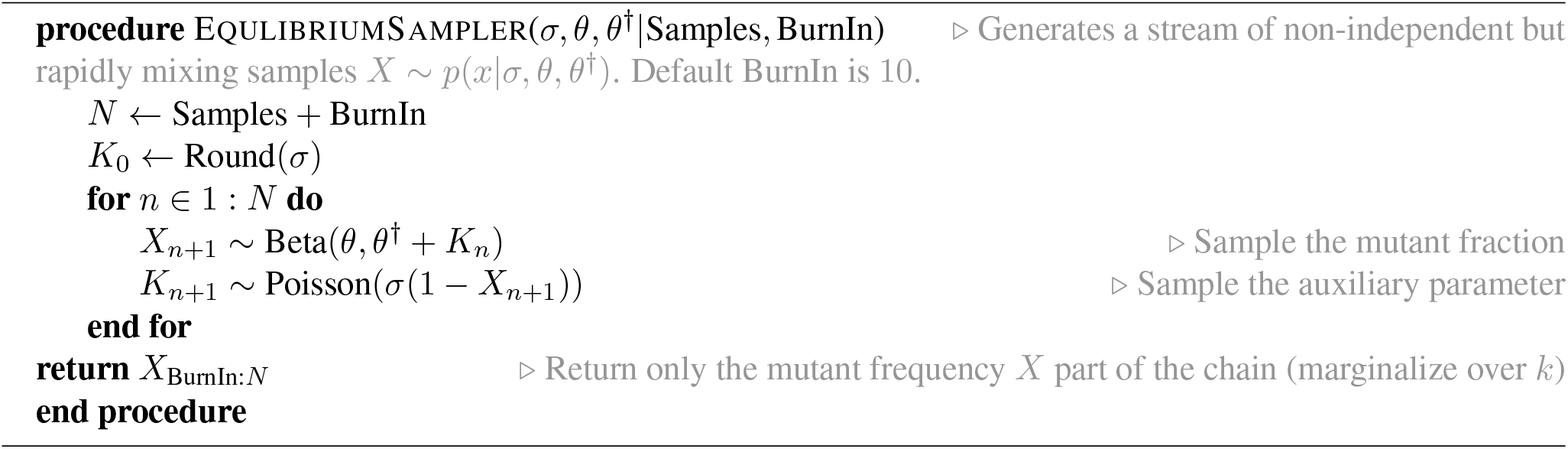

#### Simulation of the evolutionary process

We use a Gillespie algorithm to simulate the evolutionary process, where we break up the reaction calculation into two parts: randomly choosing a viral strain *ρ*_*i*_ from the population and then determining whether it reproduces or dies based on its fitness *f*_*i*_ and escape status (Algorithm 3).

### 3.5 Determining the simulation parameters of the birth-death process from genetic data

We set the intrinsic growth rate (fitness) of the wild-type virus, in the absence of competition to be *γ* = (3 days)^*−*1^, consistent with intra-patient doubling time of the virus [13, 14, 22]. We infer the neutralization rate *r* by fitting the viremia curves Fig. S1 in the trials under study, and use the averaged decay rate *r* = 0.31 for simulations, fitted using eq. S5. For the absolute mutation rate *µ*_ts_ (per nucleotide per day) we use 1.1 ×10^−5^ which is the average of the reported values for transitions per site per day from ref. [23]. Using the covariance of neutral diversity in two-fold and four-fold synonymous sites, we determine the transition/transversion diversity ratio to be *θ*_ts_*/θ*_tv_ = 7.8 (Fig. S4 and Section 4.1). We use these values to determine the forward and backward mutation rates *µ*_*s*_ and 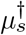 for each site (Section 4.2).

#### Algorithm 3

Population time step

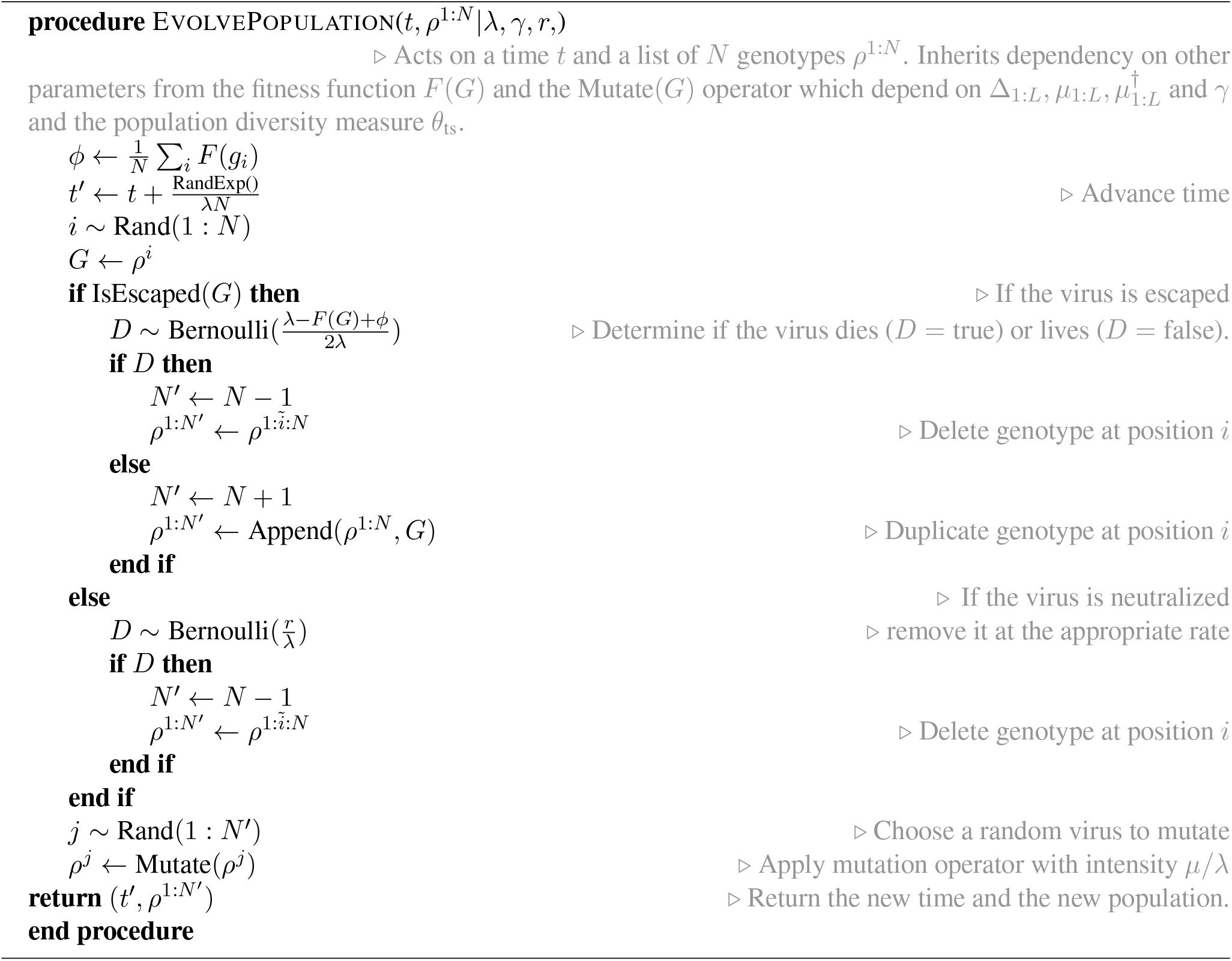

Generally the trial patients show a larger viral diversity at the start of the trial compared to the patients enrolled in the high-throughput study of ref. [4] (Fig. 2). We account for differences in the genetic makeup of the patients enrolled in the trial by directly estimating the viral diversity *θ*_ts_ from the neutral site-frequency data of patients, before the start of the trial. The estimated viral diversity *θ*_ts_, coupled with the mutation rate *µ* = 1.1×10^−5^ /day / nt, and the total rate of birth and death events in the viral population *λ*, set the carrying capacity *N*_*k*_ for a given individual,

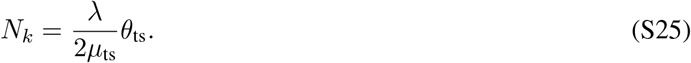

It should be noted that the value of *N*_*k*_, as the number of viruses in our individual-based simulations, is not related to the maximal viral load in the viremia measurements (i.e., steady state copy number per ml) as this relationship depends on the microscopic details of the population dynamics.

The total rate of birth and death events *λ* = *β* + *δ* tunes the amount of stochasticity, i.e., more events cause noisier dynamics. Notably, stochasticity can be linked to the size of the population *N*_*k*_, which is directly coupled to *λ* (eq. S25). We set the value of *λ* self-consistently by requiring that the minimum frequency of a variant in our simulations 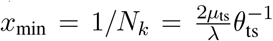 to be smaller than the escape threshold 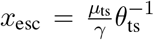 due to stochasticity (eq. S21). We set *λ* = 2day^*−*1^ so that 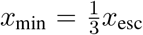. Increasing *λ* results in an in crease in the size of population *N*_*k*_ in our simulations, which is computationally costly, without qualitatively changing the statistics of the rebound trajectories.

## 4 Inference of diversity, mutational target size, and selection from genetic data

### 4.1 Inference of mutation rates and the neutral diversity within a population

Previous work has indicated an order of magnitude difference between the rate of transitions (mutations within a nucleotide class) and transversions (out-class mutations) in HIV [23–26]. Therefore, to infer the neutral diversity parameter *θ*_ts_, we also account for the differences between transition and transversion rates.

Consider the set of sequences sampled from a patient’s viral population at a particular time. Two neutral alleles that are linked by a symmetric mutational process *µ*_1*→*2_ = *µ*_2*→*1_ have a simple count likelihood. The probability to see allele 1 with multiplicity *n* and allele 2 with multiplicity *m* is given by a binomial distribution Binom(*n*, |*m x*) with parameter *x* denoting the probability for occurrence of allele 1, convolved with the neutral biallelic frequency distribution *P*_eq_(*x* | *σ* = 0, *θ*) from eq. S16. Using this probability distribution, we can evaluate the log-likelihood ℒ (*θ* |*n, m*) for the neutral diversity *θ* given the observations (*n, m*) for the multiplicities of the two alleles in the population,

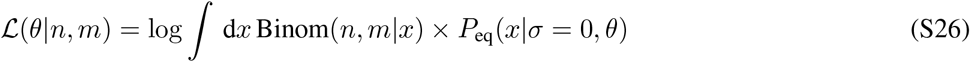

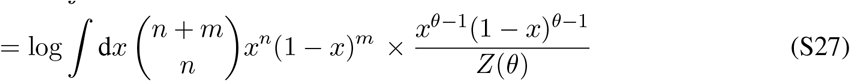

To estimate the transition diversity, we only use two-fold synonymous sites, and treat each site independently but with a shared diversity parameter *θ*_ts_. For example, consider neutral variations for two amino acids glutamine and phenylalanine. The third position in a codon for both of these amino acids are two-fold synonymous, as the two possible codons for glutamine are CAG and CAA, and for phenylalanine are TTT, and TTC. Now consider that in the data a conserved glutamine has *n* = 3 G’s and *m* = 97 A’s in the third codon position, and a conserved phenylalanine has *n* = 10 T’s and *m* = 90 C”s at its third codon position. In this case, the combined log-likelihood for the shared diversity parameter is ℒ (*θ*| data) =ℒ (*θ*| 3, 97) + ℒ (*θ* |10, 90). Extending to all of sites in the *env* protein, the maximum-likelihood estimator for the transition diversity *θ*_ts_ can be evaluated by maximizing the likelihood summed over all conserved two-fold synonymous sites,

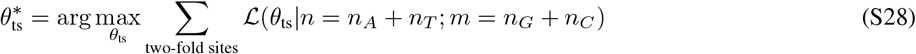

In Fig. S4 (panel A) we show that the maximum likelihood estimation method described above has better properties than the more commonly used estimator of the variance 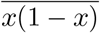 [27].

In a similar way, the likelihood for the transversion *θ*_tv_ is determined from polymorphic data at all conserved four-fold synonymous sites. One such example is the third position in a glycine codon, where (GGT, GGC, GGA, GGG) translate to the same amino acid. The maximum-likelihood estimator for the transversions is

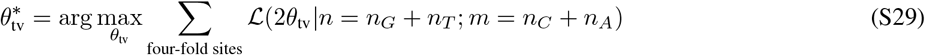

The factor of 2 in the argument of the likelihood accounts for the multiplicity of mutational pathways, e.g. from a *G* nucleotide there are two transversion possibilities, *G* →*C* and *G*→ *A* for moving from one allele to the other [28].

Using this likelihood approach, we can infer the neutral diversities 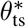 and 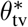 for each patient at each time point from the polymorphism in two-fold and four-fold synonymous sites. To characterize the ratio of transition to transversion rates, we use linear regression on the entire patient population and sample history and infer a constant ratio *µ*_ts_*/µ*_tv_ = *θ*_ts_*/θ*_tv_ = 7.8 (Fig. S4B). In Fig. S4, we also show that the estimate for this ratio is relatively consistent across different data sources, produced even by different sequencing technologies. The previously reported relative rate of transitions to transversions, based on the estimates of sequence divergence along phylogenetic trees of HIV-1 is *µ*_ts_*/µ*_tv_ = 5.6 [23], which is similar to our maximum likelihood estimate.

### 4.2 Inference of mutational target size for each bNAb

The nucleotide triplets which encode for amino acids at an escape site undergoe substitutions which can change the amino acid type and create an escape variant. The changes in the state of an amino acid codon can be modeled as a Markov jump process and can be visualized as a weighted graph where the nodes represent codon states, and edges represent single nucleotide substitutions linking two codon states (Fig. 1D). In our mutational model, these edges have weights associated with either the mutation rates for transitions *µ*_ts_ or transversions *µ*_tv_. We call this the *codon substitution graph*.

The codon states can be clustered into three distinct classes: (i) codons which are fatal *F*, (ii) wild-type (i.e., susceptible to neutralization by the bNAb) *W*, and escape mutants (i.e. resistant to the bNAb) *M*. We expect the escape mutants to be at a selective disadvantage compared to the resistant wild-type, and that the most common escape codons to be those which are adjacent to wild-type states.

The mutational target size is determined by the density of paths from the wild-type *W* to the escape mutants *M*.

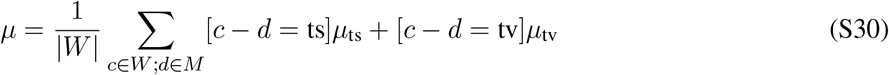

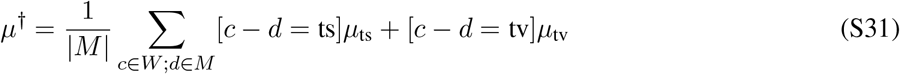

The functions [*c −d* = ts] or [*c −d* = tv] are 1 when the two codons are separated by a transition or transversion, and are zero otherwise. Note that since we only have an estimate for the ratio of the transition to transversion rates *µ*_tv_*/µ*_ts_, we can only determine the scaled mutational target sizes, 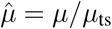 and 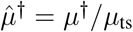, which are sufficient for inference of selection in the next section. The full list of mutational target sizes inferred for the bNAbs in this study are shown in Fig. S1.

When discussing the mutational target size of escape from a given bNAb, we refer to the total mutation rate from the susceptible (wild type) to the escape variant as,

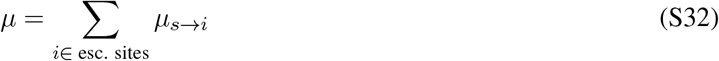

where the sum runs over all the mediating escape sites *i*, and can be interpreted as the average number of accessible escape variants. In the strong selection regime, we can write the frequency of escape mutants as,

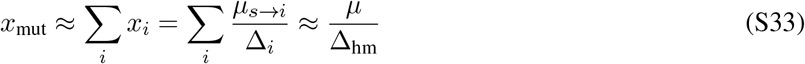

### 4.3 Inference of selection for escape mutations against each bNAb

Here, we develop an approximate likelihood approach to infer the selection ratio 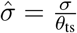, using the high-throughput sequence data from bNAb-naive HIV patients from ref. [23]. The quantity 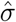 is a dimensionless ratio which is independent of the coalescence timescale *N*_*e*_, and therefore, represents a stable target for inference.

We assume that the probability to sample *m* escape mutants and *s* susceptible (wild-type) alleles at a given site in the genome of HIV in a population sampled from a patient at a given time point follows a binomial distribution Binom(*m, s* |*x*), governed by the underlying frequency *x* of the mutant allele. In addition, the frequency *x* of the allele of interest itself is drawn from the equilibrium distribution *P*_eq_(*x* |*σ, θ, θ*^*†*^) (eq. S16), governed by the diversity *θ*_ts_ inferred from the neutral sites, the estimated mutational target sizes 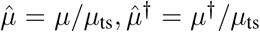, and the unknown selection ratio 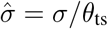. As a result, we can characterize the probability 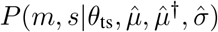 to sample *m* escape mutants and *s* susceptible-type alleles, given the scaled selection and diversity parameters as,

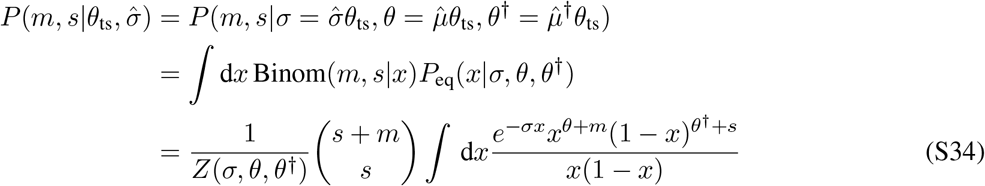

Here, *Z*(*σ, θ, θ*^*†*^) is a confluent hypergeometric function of the model parameters that sets the normalization factor for the allele frequency distribution *P*_eq_(*x*) (eq. S17). It should be noted that the viral population is in fact out of equilibrium, due to constant changes in immune pressure evolution of the B-cell and T-cell populations. Although we are ignoring these significant complications, we later use the same equilibrium distribution in a consistent way to generate standing variation in simulations. For the model to make accurate predictions, it is not necessary that the equilibrium model be exactly correct, but only that it is rich enough to provide a consistent description for the distribution of mutant frequencies observed across viral populations.

We will use the probability density in eq. S34 to define a log-likelihood function in order to infer the scaled selection 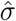 from data. To do so, we first express the logarithm of this probability density as,

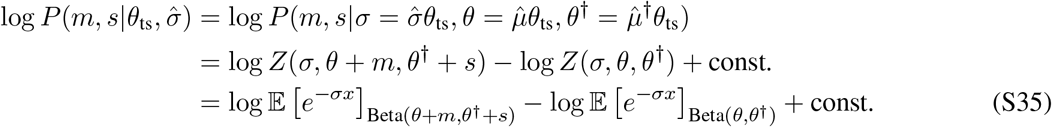

where the constant factors (const.) are independent of selection, and 𝔼 [·] _Beta(*·*)_ denotes the expectation of the argument over a Beta distribution with parameters specified in the subscript. The expression in eq. S35 implies that we can evaluate the likelihood of selection strength by computing the difference between the logarithms of the expectation for *e*^*−σx*^ over allele frequencies drawn from two neutral distributions (Beta distributions), with parameters (*θ, θ*^*†*^) and (*θ* + *m, θ*^*†*^ + *s*), respectively. This approach is more attractive as it would not require direct evaluation of the confluent hypergeometric functions for the normalization factors in eq. S34. Estimating these normalization factors is computationally intensive for large values of *σ*, since many terms in the underlying hypergeometric series should be taken into account to stably compute them. However, evaluating the expectations via sampling from these two neutral distributions has the disadvantage that it is subject to variations across simulations. We reduce the variance of our estimate of log *P* (*m, s* |*σ, θ, θ*^*†*^) in eq. S35 by using a mixture-importance sampling scheme [29] with the details shown in Algorithm 4.

We use data collected across all time points and from all patients to infer reliable estimates for selection strengths. However, allele frequencies are correlated across time points within patients (Fig. 2B), and thus, sequential measurements are not independent data points. Our estimates indicate a coalescence time of about *N*_*e*_ ∼10^3^ days based on the estimates for the mutation rate *µ*_ts_ = 10^−5^ /nt/day, and the neutral diversity *θ*_ts_ = 2*N*_*e*_*µ*_ts_ = 0.01.

#### Algorithm 4

Importance sampled log-likelihood given a single datapoint

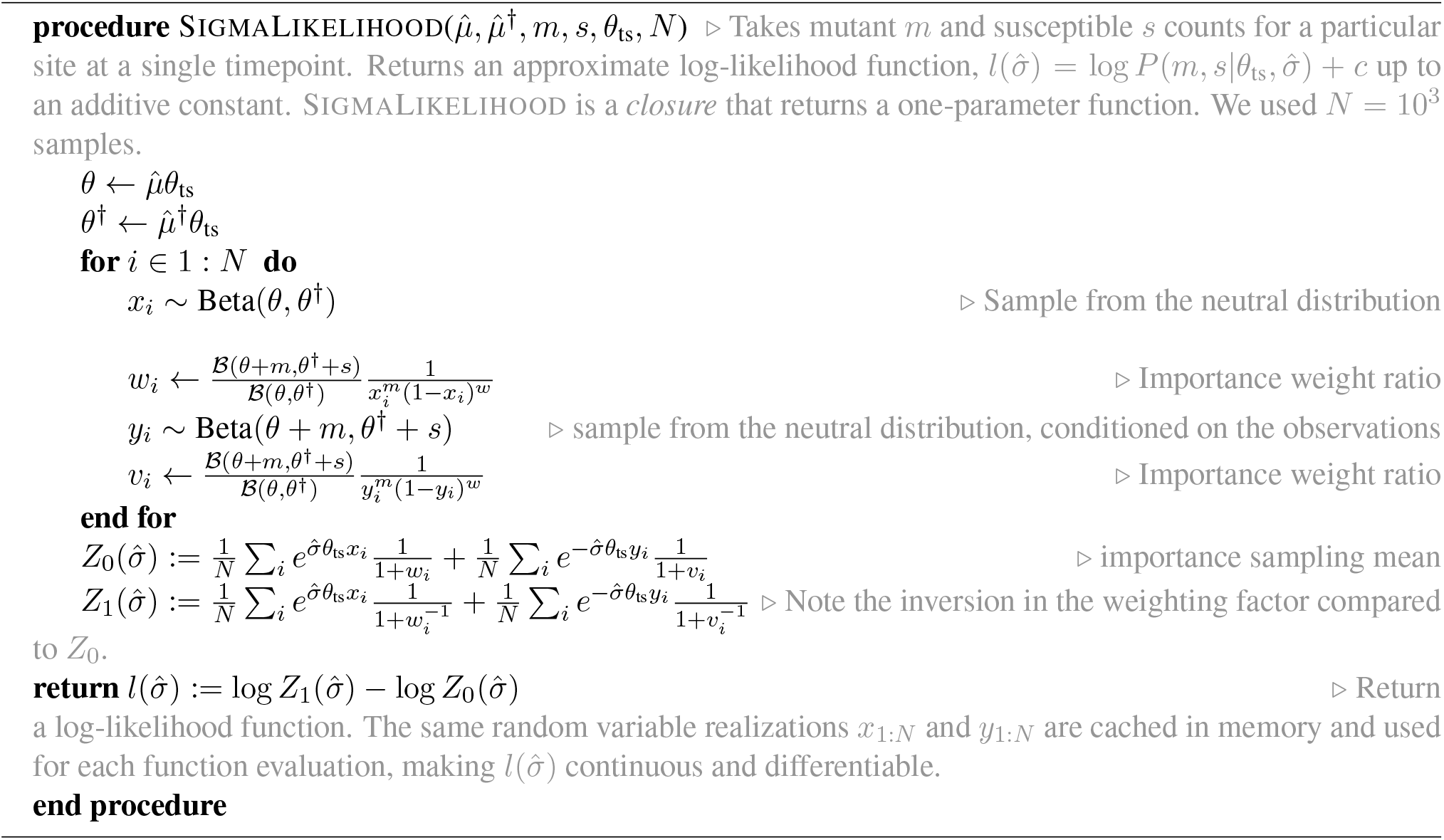

This coalescence time is much longer that the typical separation between sampled time points within a patient (∼10^2^ days), suggesting that sequential samples collected from each individual in this data are correlated. Therefore, we treat each patient as effectively a single observation, using the time-averaged likelihood for the (scaled) selection factor 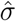:

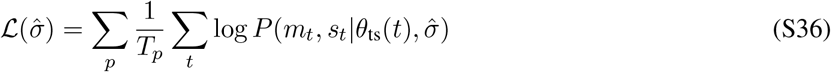

where *p* and *t* denote patient identity and sampled time points, respectively, and *T*_*p*_ is the total number of time points sampled in patient *p*. We use the likelihood in eq. S36 to generate samples from the posterior distribution for selection strengths under a flat prior, with a standard Metropolis-Hastings algorithm [30]. Since the prior is constant, this procedure amounts to simply accepting or rejecting samples based on the likelihood ratio of eq. S36. We used the centered-normal distribution with standard deviation of 50 (×*µ*_ts_ in absolute units) as the proposal density for the jumps in the Markov chain.

## 5 Predicting trial outcomes from genetically informed evolutionary models

### Predicting rebound times

We expect different distributions of patient outcomes depending on whether they have been recently infected and thus have relatively low viral diversity, or whether their infection is longstanding with a diverse viral population. To construct the distribution of initial population diversities *θ*_ts_ for simulating trial outcomes, we apply the *θ*_ts_ inference procedure (eq. S26) to pre-treatment sequence datasets available from the clinical trials under consideration. In Fig. 2D this set of pre-trial *θ*_ts_ is compared to the longitudinal in-patient diversity. We used random draws from the inferred *θ*_ts_ values for patients to generate *θ*_ts_ for simulations.

We found that there was considerably more viral escape and non-responders in our simulations than in the observed data as shown in Fig. S7A. This is *in addition* to the fact that the patients were screened to have only susceptible variants to the antibodies used in trials [1–3]. In theory, there should be zero non-responders, as such patients should have been excluded by screening. The over-prediction of *both* non-responders and late rebounds is a signature of undercounting the effective diversity of the viral populations.

The failure of both screening and our naive prediction in undercounting the diversity in the viral population can be explained by an effective viral reservoir. Viral variants which mediate rebound can come from compartments such as including bone marrow, lymph nodes, and organ tissues, and can be genetically distinct from those sample from the plasma T-cells during screening [31–33]. This reservoir of viral diversity can reappear in plasma after infusion of a bNAb and could in part contribute to treatment failure [34–37].

### Determining patient diversity enhancement due to latent reservoirs

We model the effect of reservoirs as a simple inflation of the diversity observed by a multiplicative factor *ξ*. We fit *ξ* directly to trial observations, using a disparity-based approach by minimizing an empirical divergence estimator [38] between the observed and simulated data. To do so, we characterize the Hellinger distance [39, 40] between the true distribution of rebound times *P* (*t*) and the rebound times *Q*(*t*|*ξ*) generated by simulations with a given reservoir factor *ξ*,

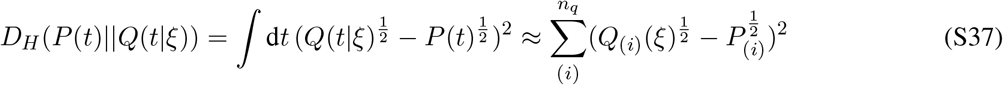

Algorithm 5 defines the procedure that we use to estimate the Hellinger distance *D*_*H*_ (*Q*(*t*)||*P* (*t*|*ξ*)). Specifically, we use *n*_*q*_ quantiles of the observed data *x*_(*i*)_ ∼ *Q* to partition the space of observations into discrete outcomes

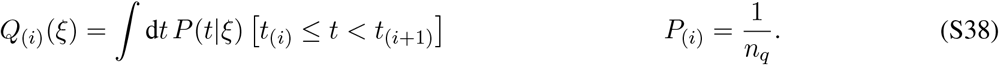

where *P*_(*i*)_ is a constant by construction, and *P*_(*i*)_(*ξ*) is estimated by simulations, and [·] is the Iverson bracket [41] (Algorithm 5)

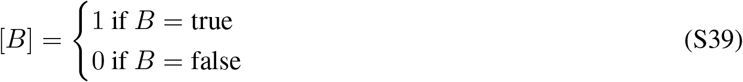

To simulate data for this analysis, we generate *S* rebound times (*T*_1:*S*_) by simulations, given the scaled diversity values *ξθ*_ts_. We then find the optimal value *ξ*^*∗*^ by minimizing the disparity with the observed rebound times *t*_1:*p*_ by brute-force search,

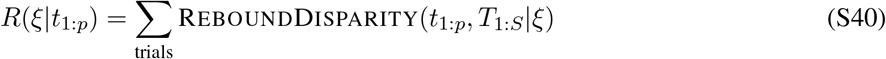

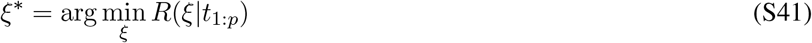

Here, REBOUNDDISPARITY is the function defined by Algorithm 5; see ref. [38] for details. We find the optimal reservoir factor to be *ξ*^*∗*^ = 2.1, which we use in subsequent therapy prediction. The disparity over various values of *ξ* for different trials is shown in Fig. S7.

### Simulating outcomes of the clinal trails

Given the reservoir-corrected estimate of the diversity *ξθ* and the posterior samples for selection factors 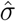, we now summarize how we simulated the outcome of clinical trials.

For a given bNAb, we draw the selection factor at each of the escape mediating sites from the corresponding Bayesian posterior on 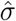; the posterior distributions are shown in Fig. 4A, and summarized in Table S1. We also use the mutational target size (forward *µ* and backward *µ*^*†*^ rates) associated with each of the escape mediating sites of a given bNAb; see Table S1. The result can be summarized in a mutation / selection matrix 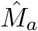 for a given bNAb *a*, where each column corresponds to an escape mediating site *i* against the bNAb,

#### Algorithm 5

Rebound-time Disparity

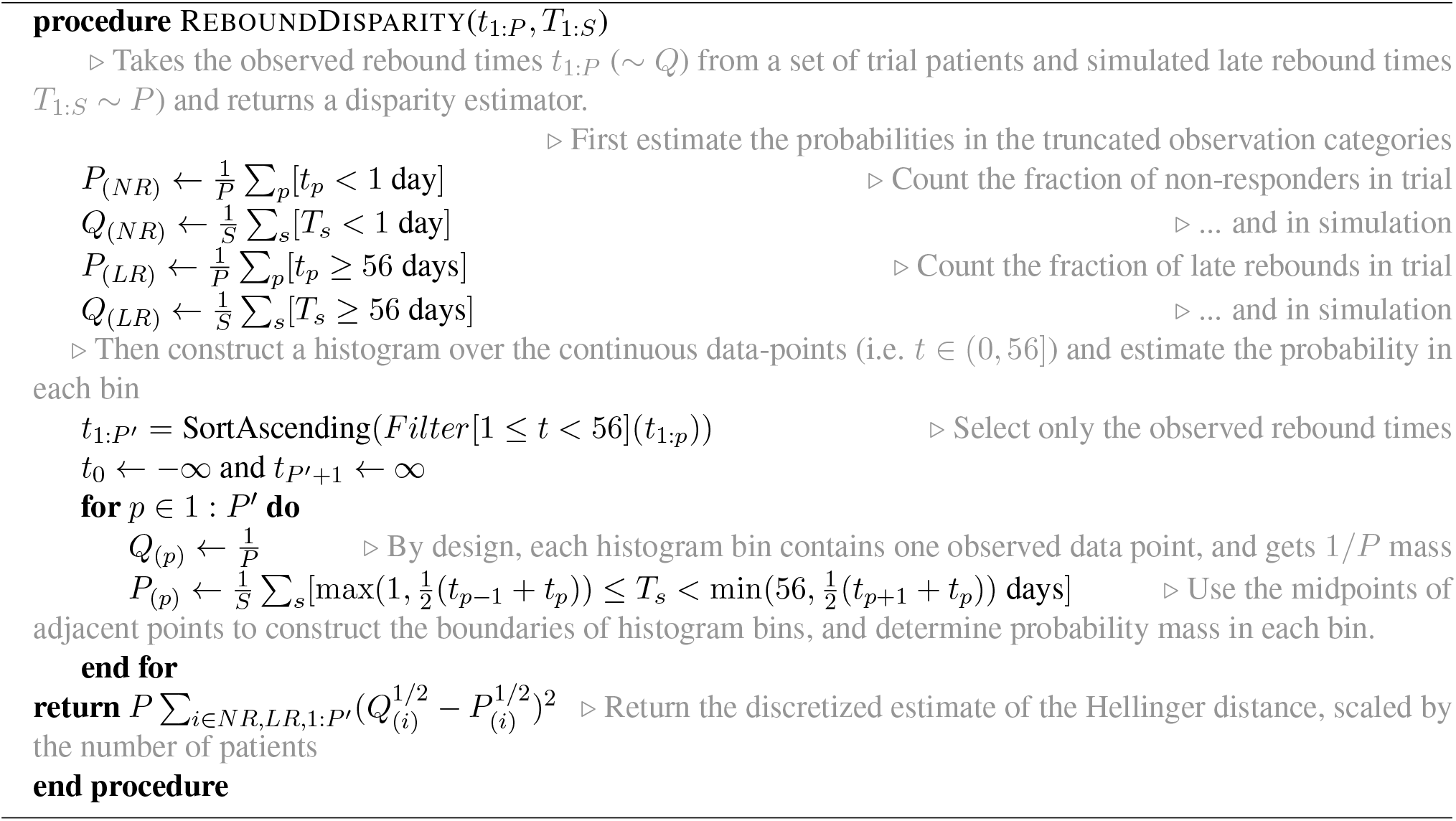

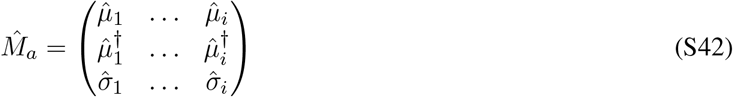

The elements of the matrix 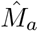 are the scaled mutation and selection factors, i.e., 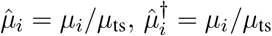, and 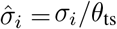, where the absolute value of mutation rate is set to *µ*_ts_ = 1.11 × 10^−5^ */*nt*/*day from ref. [23].

For each patient in our simulated trial, we then draw diversity *θ*_ts_ from the patient pool, and scale it by our fitted *ξ* = 2.1, resulting in patient-specific selection and mutation factors,

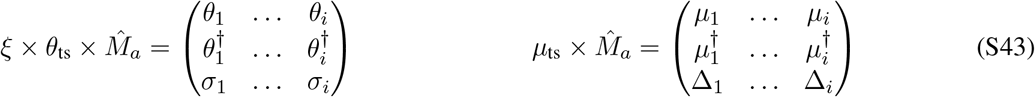

These parameters are then used to initialize the state of an HIV population within a patient according to Algorithm 1, and to determine the absolute rates in Algorithm 3 for the population evolution according to eq. S43. The decay rate is set to the fitted trial average of *r* = 0.31 days^*−*1^ (eq. S5). The carrying capacity *N*_*k*_ is set according to eq. S25. This determines all parameters of the birth-death process simulating the intra-patient evolution of HIV, which are used in Algorithm 3.

We evolve a population through time until 56 days have elapsed since treatment, or until the escape fraction relative to the carrying capacity *x*_*t*_ is above 0.8 (Algorithm 3). After *x*_*t*_ *>*. 8 the evolution is governed by the deterministic equations, and the stochastic simulation ends. The rebound time *T*, defined as the intersection of the exponential envelope and the carrying capacity, can then be calculated analytically as,

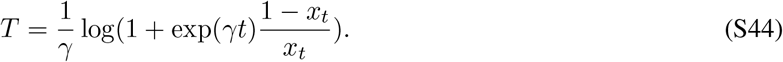

The resulting distribution for rebound times are shown as model predictions in Fig. 3B-D.

Rebound times generated in this fashion were also used to estimate the probability of late rebound to characterize the efficacy of a given bNAb in curbing viral rebound. The probability of late rebound was estimated from 10^4^ simulated patients. The interdecile quantiles (0.1 - 0.9) of early rebound probability over 200 values of scaled selection coefficients 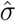drawn from the posteriors in Fig. 4A are shown in Fig. 4C.

## 6 Model robustness

### Effect of genomic linkage on the inference of selection

In our inference of selection (eq. S36, Fig. 4A), we assume that the escape-mediating sites are at linkage equilibrium and that the distribution of allele frequencies can be approximated by a skewed Beta distribution (eq. S16), reflecting the equilibrium of allele frequencies. In reality, despite recombination, the HIV genome exhibits linkage effects, especially at nearby sites [4], and the viral populations experience changing selective pressures by the immune system [25,42,43], and the transient population bottlenecks during therapy [44].

To test the limits on the validity of our inference procedure, we applied it to *in-silico* populations generated by full-genome forward-time simulations (Algorithm 3) in the presence and absence of recombination. To do so, we considered an ensemble of ten patients with 100 genomes sampled at ten time points, and used two diversity parameters *θ*_ts_ = 0.01 and *θ*_ts_ = 0.1, to cover the range reflected in patient data (Fig. 2D, S6).

One relevant scenario to consider is the impact of other selected sites in the genome on the distribution of alleles at the escape mediating sites against bNAbs. The sites under a strong constant selection are likely to be already fixed (or at a high a frequency) at their favorable state in the population. However, the strong selection on a large fraction of antigenic sites in HIV can be thought as time-varying, due to the changing pressure imposed by the immune system or therapy. To capture this effect, we simulated whole genome evolution in which linked sites were under strong selection (0.1 × growth rate), and where the sign of selection changed after exponentially distributed waiting times (i.e. as a Poisson process); this model of fluctuating selection has been used in the context of influenza evolution [45], and for somatic evolution of B-cell repertoires in HIV patients [43]. The resulted evolutionary dynamics in this case can involve strong selective sweeps and clonal interference due to the continuous rise of beneficial mutations (in the linked sites) within a population.

To test the robustness of our selection inference, we evaluated the distribution of maximum likelihood estimates (MLEs) for the selection values 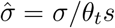 at the escape mediating sites, inferred from the ensemble of sequences obtain from simulated data with linkage. Fig. S6 shows that even for fully linked genomes (zero recombination) our MLE estimate of selection has little bias relative to the true values used in the simulations. Adding recombination into the simulations only further attenuates the effect of linkage (Fig. S6), making the estimates more accurate.

The reason that selective sweeps of linked beneficial mutations have only minor effects on our inference of selection for the escape mediating sites is two-fold: First, the primary mechanism by which selective sweeps change the strength of selection at linked sites is via a reduction in the effective population size [46, 47]. However, variations in the effective population size are already accounted for in our inference procedure: The selection likelihood in eq. S36 is conditioned on the measured neutral-site diversity, *θ* = 2*N*_*e*_*µ*. The change in the effective population size impacts the selection coefficient *σ* = 2*N*_*e*_Δ and the diversity *θ* = 2*N*_*e*_*µ* in the same way, and therefore, the (scaled) selection parameter 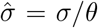 that we infer from data remains insensitive to changes in the effective population size.

The second reason for the robustness of our selection inference to linkage is due to the fact that a beneficial allele in a linked locus can appear on a genetic background with or without a susceptible variant, leading to the rise of either variants in the population. As a results, the impact of such hitchhiking remains a secondary issue in inference of selection at the escape sites, for which an ensemble of populations from different patients with distinct evolutionary histories of HIV is used.

### Robustness of selection inference to intra-patient temporal correlations among HIV strains

To infer the selection effect of mutations from the longitudinal deep sequencing data of [4], we use time averaging of the likelihood (eq. S36) to avoid conflating our results due to temporal correlations between the circulating alleles within patients (Fig. 2B). We can view this choice as being one choice among two extremes: (i) to treat each patient as effectively one independent data point so that all patients are given the same weight, or (ii) to treat each time point as independent, giving patients with more time points a higher weight. These two choices correspond to different log-likelihood functions for the (scaled) selection factor 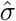:

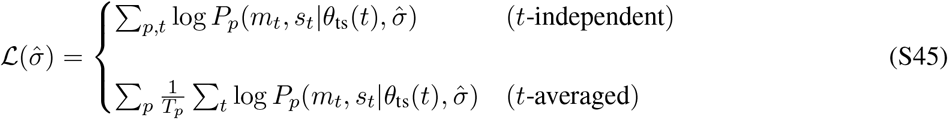

where *T*_*p*_ is the total number of time points from patient *p*, and 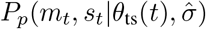 is the probability to observe *m* escape mutants, and *s* susceptible variants at time *t* in patient *p*, given the neutral diversity *θ*_ts_(*t*) and the scaled selection factor 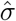.

We find that both of these approaches result in similar posteriors for selection 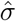 (Fig. S5A and C) although the *t*-averaged likelihood has a higher uncertainty due to fewer independent time points. Thus, our inference of selection is insensitive to the exact choice of the likelihood function given in eq. S45, yet our time-averaged approach remains the more conservative choice between the two.

### Model of viral rebound with the reservoir-corrected effects (*ξ* = 1)

In eq. S41 we introduced the reservoir factor *ξ*^*∗*^ = 2.1 to account for the diversity of HIV that is not sampled from a patient’s plasma prior to therapy,which resulted in a better fit of the rebound time distributions (Fig. 3) compared to a reservoir-free model (Fig. S7A). Here, we quantify the importance of the reservoir factor with a statistical test on the null hypothesis, *ξ*_0_ = 1. Specifically, we perform a hypothesis test to test the necessity of using an inflated diversity *ξ θ*_ts_ relative to using the bare diversity observed in pre-trial sequence data *θ*_ts_. To do so, we construct a disparity-based test statistic [38], which is analogous to the likelihood ratio test statistic.

Recall that the optimal reservoir factor *ξ*^*∗*^ = arg min_*ξ*_ *R*(*ξ* |*t*_1:*p*_) was obtained by minimizing the disparity function *R*(*ξ* |*t*_1:*p*_) across measurements of rebound times *t*_1:*p*_ from all the *p* patients in data (eq. S41). We can estimate the test statistic for the reduction in disparity between the null hypothesis, *ξ*_0_ = 1 and the fitted reservoir factor *ξ*^*∗*^ as,

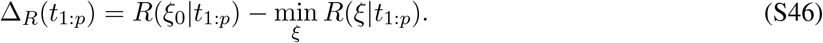

We can then determine the p-value by estimating the quantile of the observed test statistic Δ_*R*_(*t*_1:*p*_) relative to that inferred from the distribution of Δ_*R*_(*T*_1:*p*_) obtained from simulations under null hypothesis *ξ*_0_ = 1 [48]. Specifically,

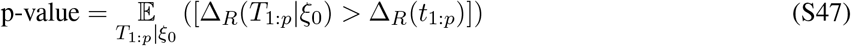

where 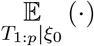 denotes the expectation over the rebound times *T*_1:*p*_ obtained from 1000 realizations of simulated populations each with *p* patients, and under the null hypothesis *ξ*_0_ = 1. [·] is the Iverson bracket that takes value 1 when its argument is true and 0, otherwise (eq. S39). The observed Δ_*R*_ (eq. S46), the distribution of simulated values of Δ_*R*_(*T*_1:*p*_|*ξ*_0_) under the null hypothesis, and the resulting p-value = 0.004 are shown in Fig. S7B,C.

It should be noted that here we use the disparity measure because the corresponding likelihood function for the reservoir factor is inaccessible through forward simulations of populations. However, a general analogy exists between our approach and the more commonly used likelihood approach. Specifically, in an analogous likelihood-ratio test, the test statistic Δ_*L*_ = max_*ξ*_ log *p*(*ξ* | *t*_1:*p*_) − log *p*(*ξ*_0_ | *t*_1:*p*_) would be asymptotically *χ*^2^-distributed with one degree of freedom under the null hypothesis [49], and the quantile under the null-hypothesis (p-value) would be estimated by inverting the *χ*^2^ cumulative distribution function (i.e. a *χ*^2^ test).

### Robustness of selection inference to strains from different clades of HIV

The longitudinal deep sequencing data of [4] is collected from 11 patients, 9 of whom are infected with clade B strains of HIV-1, which is the dominant clade circulating in Europe [50]. All of the clinical trials we considered [1–3] are from patients carrying clade B strains. For the results presented in the main text, we included all the 11 patients in our analysis. Here, we test wether our inference of selection is sensitive to the choice of including or excluding non-clade B patients in our analysis. We therefore repeated our inference procedure for selection by excluding the two non-clade B patients. Fig. S5A,B shows a strong agreement between the Bayesian posterior for selection factors in the two cases, with a slight increase in uncertainty for the case with only clade B patients. This increased uncertainty is related to the reduction in sample size by excluding the non-clade B patients from data. Nonetheless, the richness of the intra-patient diversity makes the inference robust to the exclusion of one or two patients.

### Robustness of predictions for trial efficacy to the inferred values of selection strength

How sensitive are the outcomes of our predictions for the rebound time distributions (Fig. 1) to the exact values of inferred selection strengths we used for our simulations? We addressed this question by performing a disparity analysis similar to that for the diversity *θ* in eq. S46. Specifically, we assessed whether we might need to rescale our inferred selection strength *σ/θ*_ts_ by a multiplicative factor *ξ*_*s*_ (Fig. S7D,E).

In contrast to diversity, the reduction in disparity for adjustment of selection with a factor *ξ*_*s*_ is small (Fig. S7D) and not statistically significant (p-value = 0.49; Fig. S7E), and could be attributed to count noise. Still, we cannot discount the possibility that selection was slightly overestimated, possibly due to the effect of compensatory mutations in linked genome, the interplay between the reservoir and the inference of selection, or other biological factors. Nonetheless, in absolute terms, the null hypothesis (i.e., *ξ*_*s*_ = 1) cannot be rejected and we have no statistical justification for adding an adjustment factor for selection inferred from untreated patients.

## Supplementary Figures

**Figure S1:**
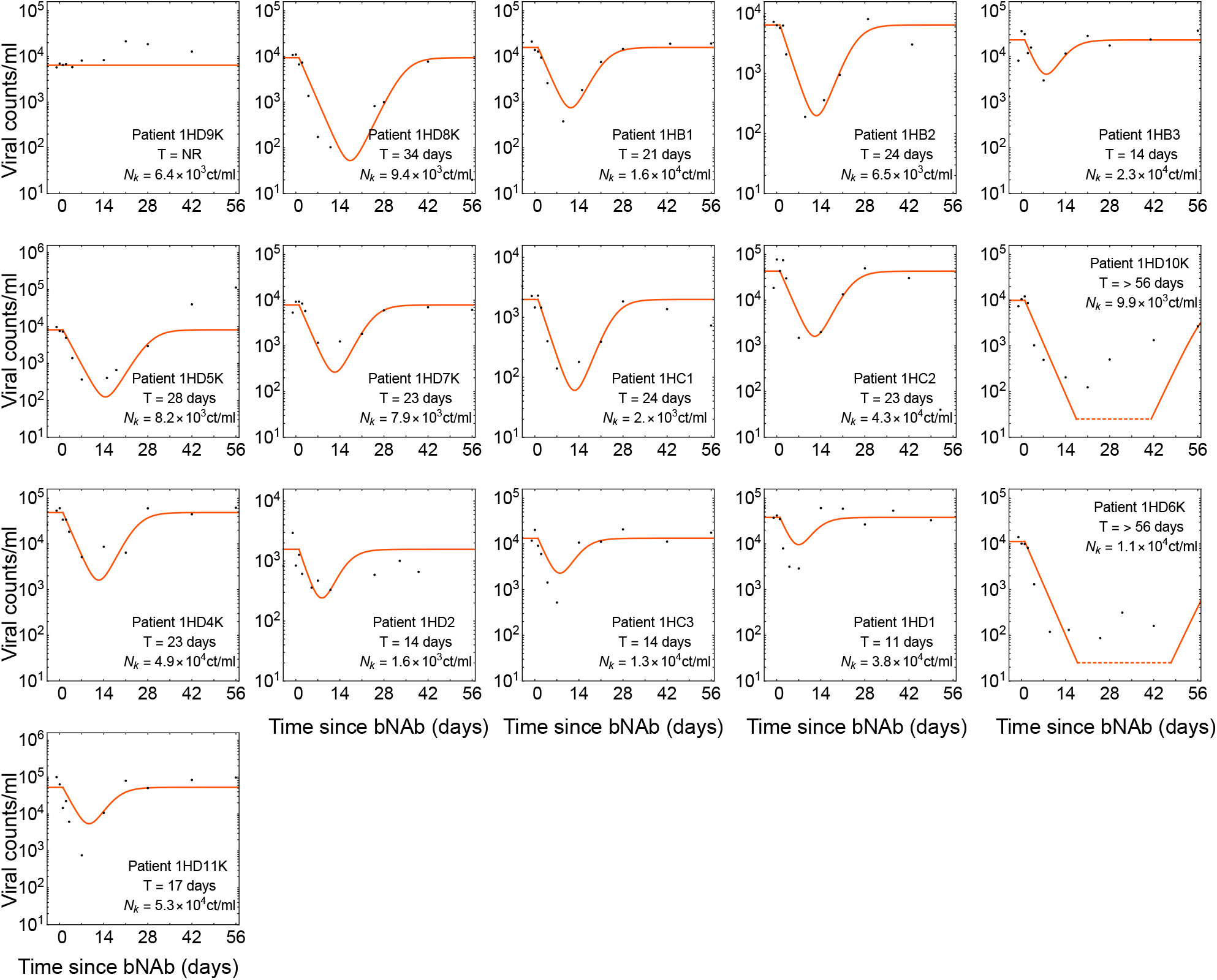
Dynamics of viremia in patients from the 10-1074 trial. Panels shows the measured viremia over time (black dots) and the fitted deterministic dynamics from eq. S2 (red line) for all the patients in the 10-1074 trial [2]. Patient-specific fitted carrying capacity *N*_*k*_ and the estimated rebound times are reported in each panel. A common decay rate *r* = 0.36 day^*−*1^ is fitted to all patient data in this trial, and growth rate is set to 0.33 day^*−*1^.

**Figure S2:**
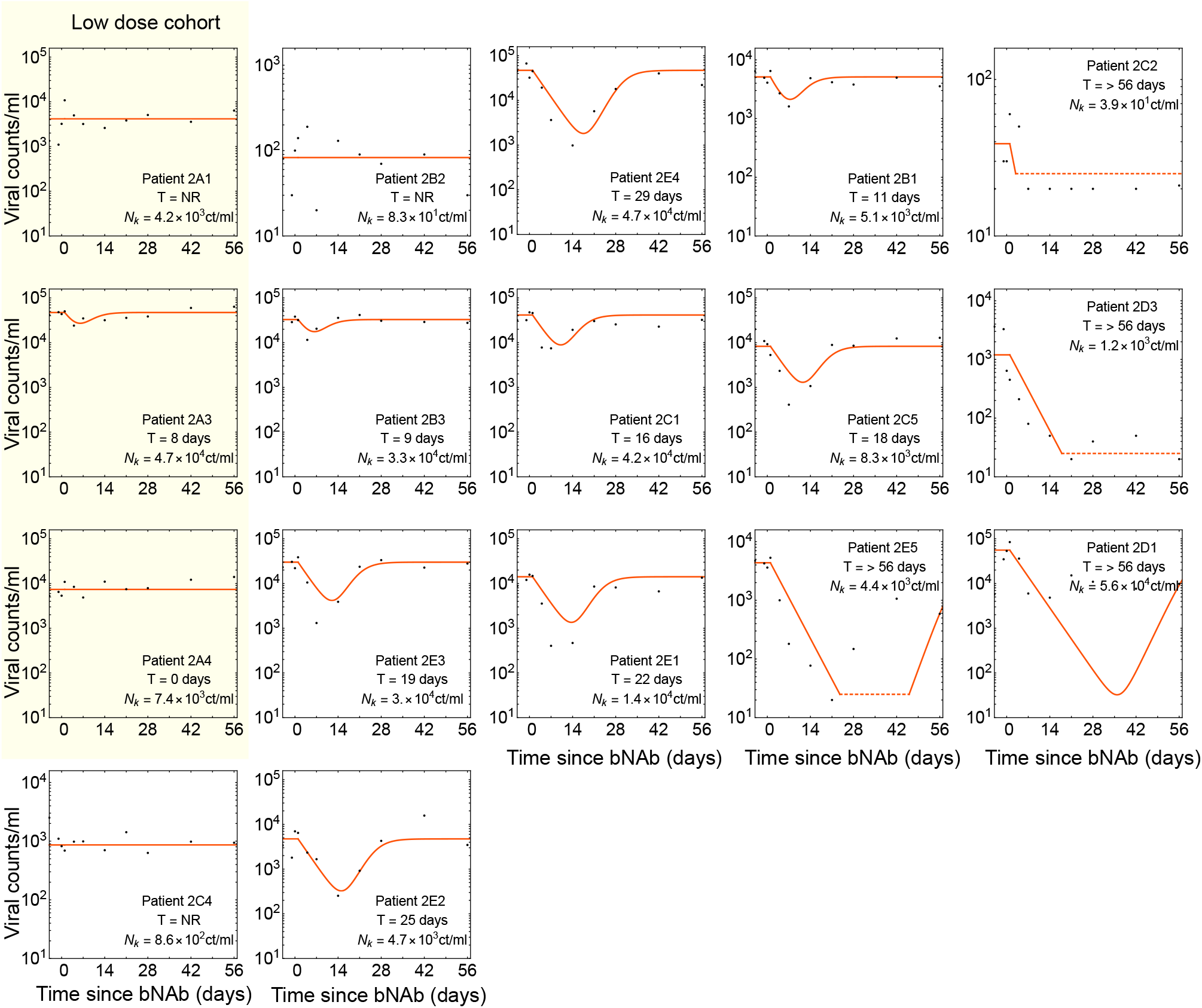
Dynamics of viremia in patients from the 3BNC117 trial. Similar to Fig. S1 but for the 3BNC117 trial [1]. The cohort treated with a low dosage of bNAb (1 mg*/*kg as opposed to 3 *−*30 mg*/*kg) is shown in yellow; these patients exhibited a very weak response to there treatment and were excluded from our analysis. A common decay rate *r* = 0.23 day^*−*1^ is fitted to all patient data in this trial, and growth rate is set to 0.33 day^*−*1^.

**Figure S3:**
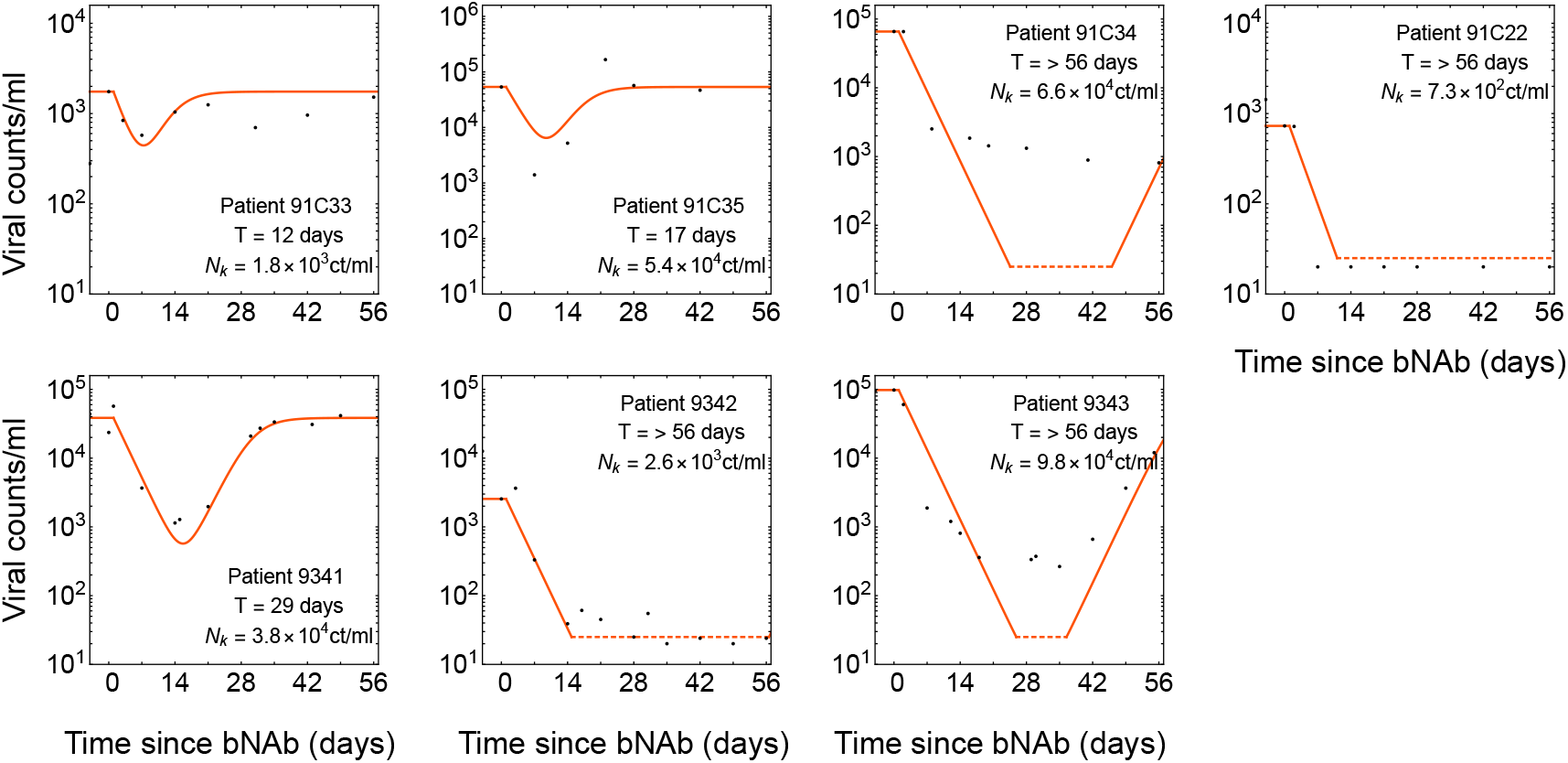
Fits to raw viremia data in combination trial. Similar to Fig. S1 but for the combination therapy with 10-1074 and 3BNC117 [3]. A common decay rate *r* = 0.33 day^*−*1^ is fitted to all patient data in this trial, and growth rate is set to 0.33 day^*−*1^.

**Figure S4:**
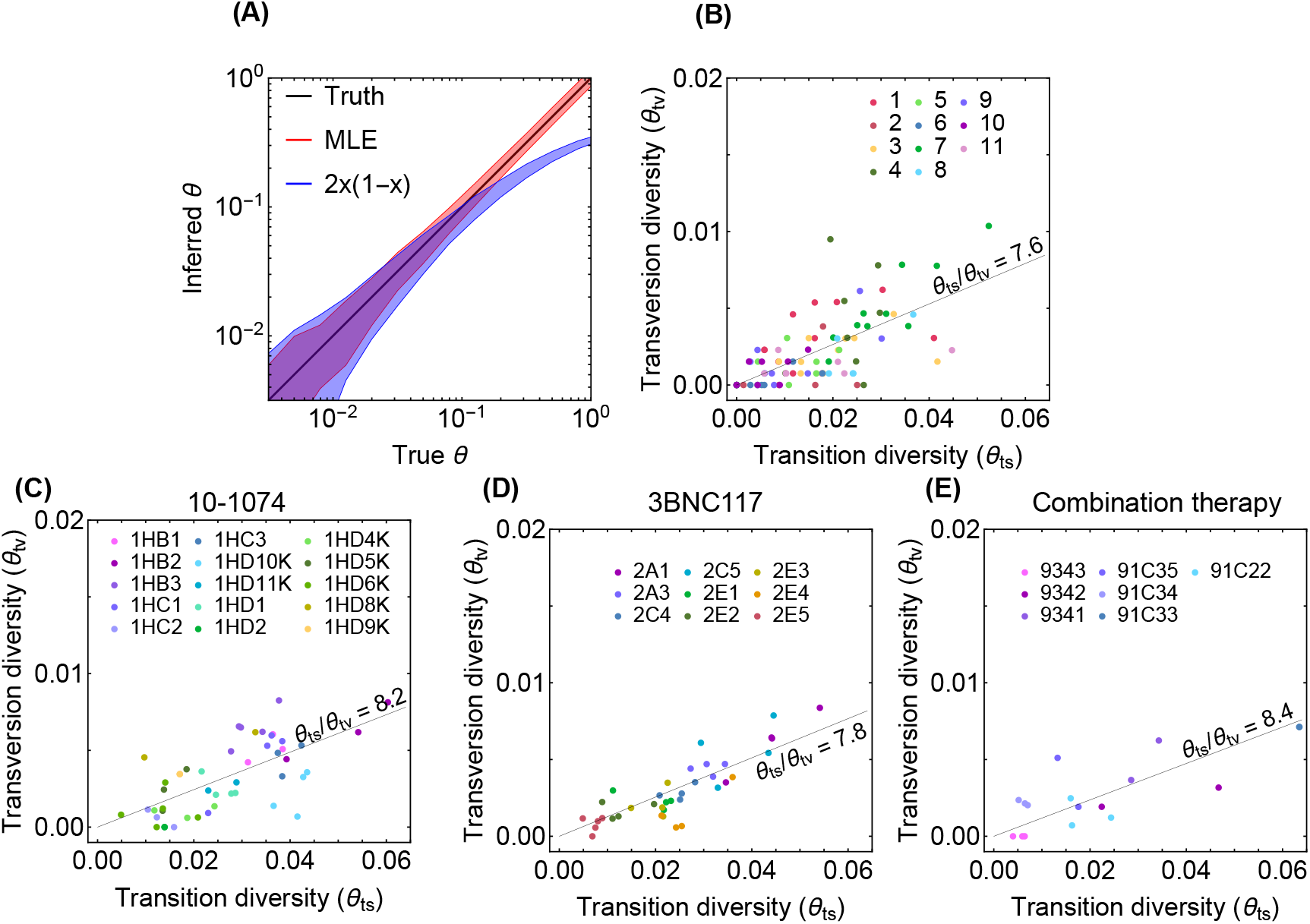
Inference of diversity from genetic data. **(A)** The inter-decile ranges of diversity estimates for 10^3^ simulations of genomes of 10^2^ independent neutral sites are shown. The true values of the diversity used for simulations are shown in black. Inter-decile maximum likelihood estimate (MLE) for the diversity *θ* from eq. S26 are shown in red, and the observed variance of allele frequency *x* as a measure of diversity *θ* = 2*x*(1 *−x*) is shown in blue. The inferred diversity associated with transversions *θ*_tv_ is shown against that of the transition *θ*_ts_ for patients (colors) enrolled in **(B)** the longitudinal study with high throughput HIV sequence data [4], and in the bNAB trials with **(C)** 10-1074 [2], **(D)** 3BNC117 [1], and **(E)** combination of 10-1074 and 3BNC117 [3]. We find that all four diversity distributions share similar ratios *θ*_ts_*/θ*_tv_, indicated in each panel.

**Figure S5:**
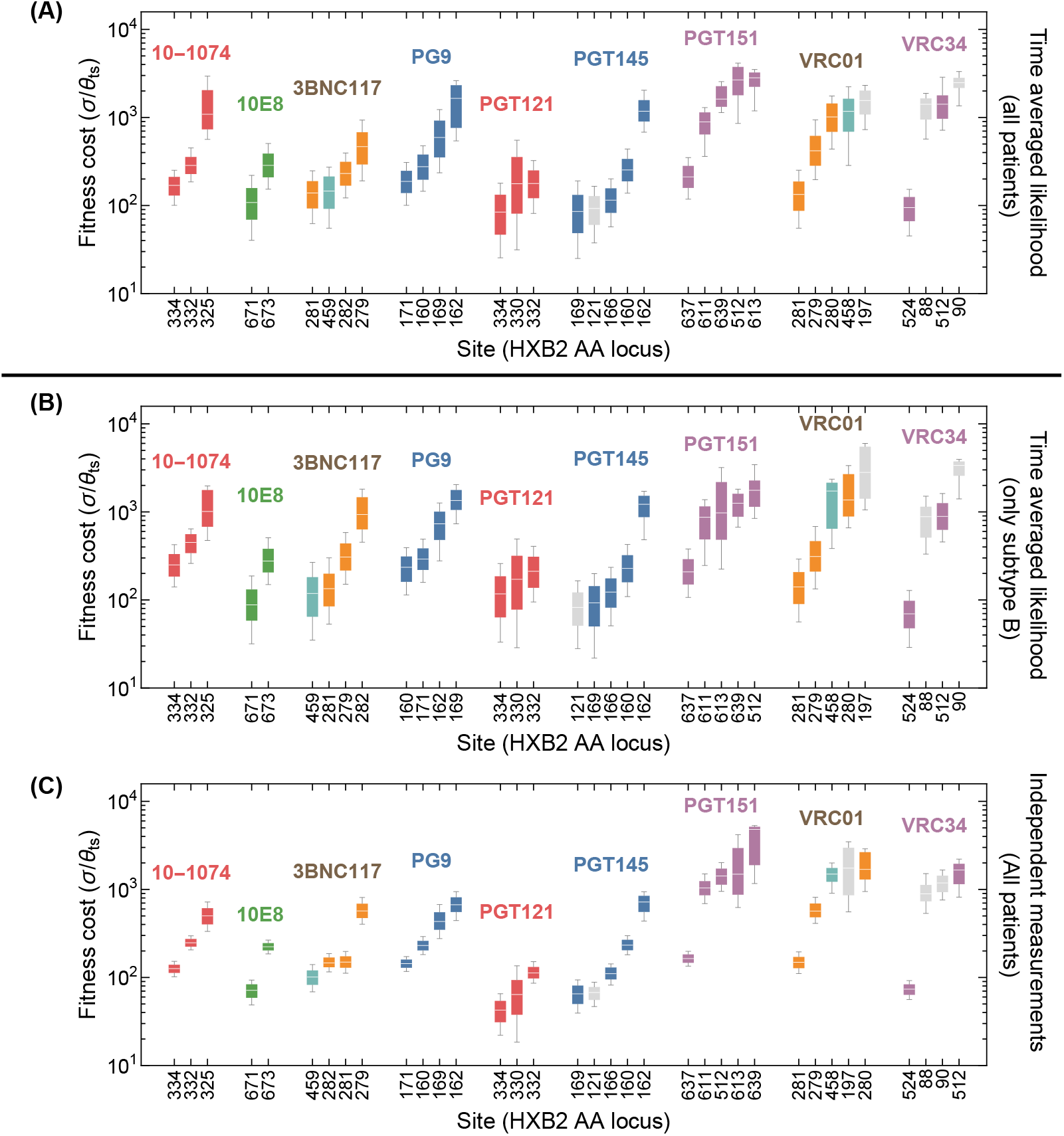
Robustness of selection inference to correlations among HIV strains. Fitness cost for escape mediating sites of different bNAbs is shown based on the selection inference using **(A)** the time-averaged likelihood (eq. S45 *t*-averaged) with data from all patients, as a reference (similar to Fig. 4A), **(B)** the time-averaged likelihood (eq. S45 *t*-averaged) with data from patients infected with HIV clade B only, and **(C)** independent time likelihood (eq. S45 *t*-independent) with data from all patients. Limiting the inference to patients infected with clade B virus in (B) result in minor changes in the inferred selection strength and a slight increase in uncertainties due to a smaller data. However, the general structure of the posteriors remain similar to (A). Treating all time points as independent in (C) increases the effective sample-size of the data and reduces model uncertainty. Nonetheless, except for slight changes on the selection ordering of escape sites against 3BNC117, the median of the selection posteriors remain similar to (A). The color code is similar to Fig. 4A and is determined by the *env* region associated with each site.

**Figure S6:**
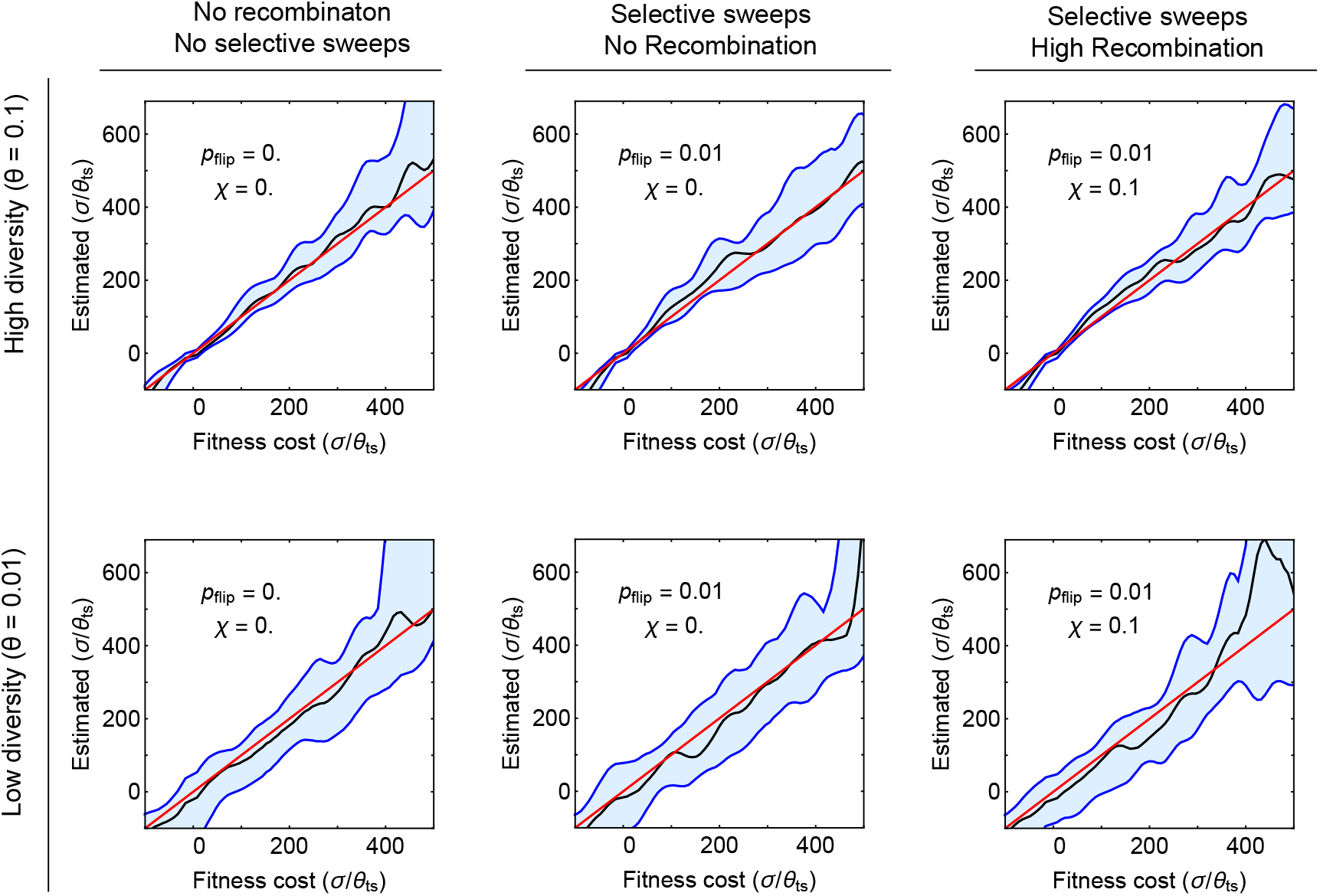
Robustness of selection inference to genetic linkage and hitchhiking. Results of selection inference for simulated populations that carry escape mediating sites linked with other selected sites in the genome are shown. The blue shading shows the interdecile ranges (0.1 -0.9 quantiles) for the inferred maximum likelihood estimates (MLE) of scaled selection 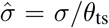 on the escape mediating sites versus the true values used in the simulations, for different genetic linkage effects (columns) and different values of population diversity (rows). Each inferred value is based on 10 realizations of simulations (*in-silico* patients) for populations evolving over time, with samples taken at 10 time points spaced at 100 days. Simulations are done on whole genomes of length 512, in which 32 sites under strong selection 0.02*f*_0_ are equally spaced throughout the genome; *f*_0_ is the base growth rate of 1.0 day^*−*1^. Each day, the preferred allele in one of the 32 sites is swapped (selection changes sign) with probability *p*_flip_. The rate of selection fluctuation *p*_flip_ and the recombination rate *χ* are given in each panel. The carrying capacity is set to *N*_*k*_ = 5, 000, and the event rate is *λ* = 2 day^*−*1^; see Algorithm 3. To initialize, we let the populations equilibrate for 4*N*_*e*_ generations, and then gather data for maximum likelihood inference of selection at the escape mediating sites. In all panels, the median MLE for selection (black line) closely matches the true selection strength (redline). Quantile lines are smoothened by a Gaussian kernel. Variation is largest when diversity is low and when mutants are rare, which concords with the results from the Bayesian inference procedure.

**Figure S7:**
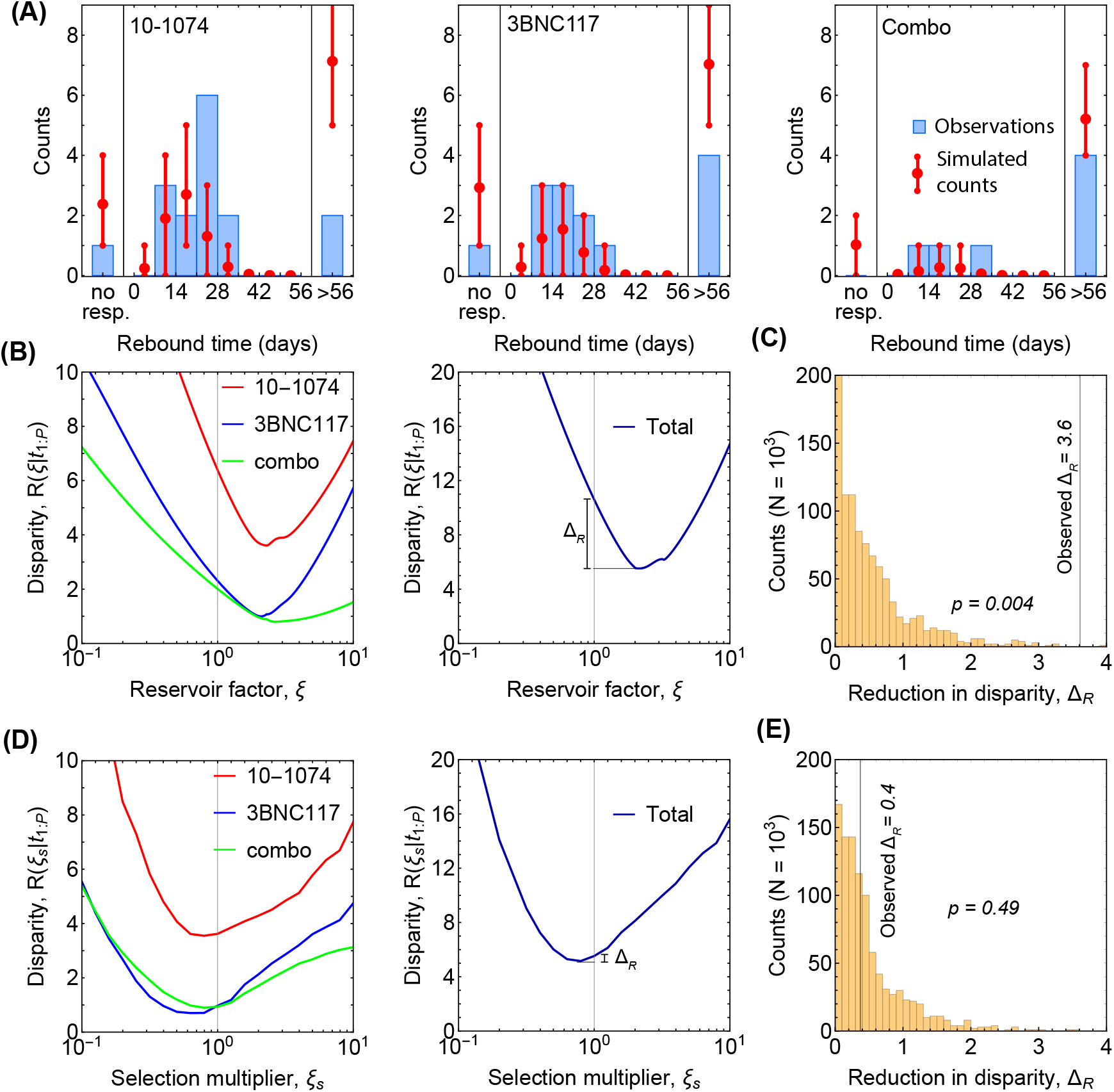
Minimum disparity estimation for adjustment of diversity and selection. **(A)** The predicted rebound times without including an increase in diversity due to reservoirs (i.e., *ξ*_0_ = 1 in eq. S47) qualitatively matches the data from the three trials (histograms in each panel), with an overestimation in the number of non-responders and patients with late rebound. For comparison, the predictions with reservoir-corrected diversity is shown in Fig. 3A. **(B)** The disparity *R*(*ξ* | *t*_1:*p*_) (eq. S40) is shown as a function of the reservoir factor *ξ* for the three trials (colors; left panel). The total disparity over all trials is shown in the right panel, where Δ_*R*_ indicates the reduction in disparity by using the optimal reservoir factor *ξ*^*∗*^ = 2.1. **(C)** The distribution for the reduction in disparity Δ_*R*_ (eq. S46) is shown for 1000 realizations of simulated data under the null hypothesis with *ξ*_0_ = 1. The large reduction in disparity indicates that the null hypothesis can be rejected with p-value = 0.004 (eq. S47). **(D, E)** Similar to (B, C) but for assessing the sensitivity of the fit to rescaling of the selection strength *σ/θ*_ts_ by a multiplicative factor *ξ*_*s*_. In this case the null hypothesis is that the inferred selection by the Bayesian procedure in eq. S36 requires no further rescaling for the model to fit the distribution of the rebound times (i.e., *ξ*_*s*_ = 1). The large p-value = 0.49 shown in (E) indicates that the null hypothesis cannot be rejected and we have no statistical justification for adding an adjustment factor for selection inferred from untreated patients.

**Table S1:** Selection and mutational target size for escape-mediating sites against each bNAb. Shown are the sites (column 1) and the susceptible and the escape amino acids (column 2) for each bNAb. We called patterns for 10-1074 and CD4bs antibodies VRC01 and 3BNC117 using genetic trial data and the remainder using DMS data. The inferred mutational target size of escape at each site (forward *µ* and backward *µ*^*†*^ mutation rates) is shown in column 3. The major quantiles (10%, 50% (median), 90%) associated with the inferred site-specific Bayesian posterior of the scaled selection strength 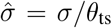 are shown in column 4. The corresponding quantiles for the strength of selection 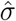, after convolving the posterior for the scaled selection 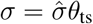 with the reservoir-corrected intra-patient diversity of HIV *ξθ* are shown in column 5.

| bNAb | site | susceptible AA | escape AA | $\mu$ $\mu^\dagger$ | | $\sigma/\theta$ quantiles | | | $\sigma$ quantiles | | |
| --- | --- | --- | --- | --- | --- | --- | --- | --- | --- | --- | --- |
| | | | | $\mu$ | $\mu^\dagger$ | 0.1 | 0.5 | 0.9 | 0.1 | 0.5 | 0.9 |
| 10-1074 | 325 | DN | EGK | 0.76 | 0.51 | $5.6 \cdot 10^2$ | $1.1 \cdot 10^3$ | $2.9 \cdot 10^3$ | 8.8 | 29 | 93 |
| | 332 | N | DHIKSTY | 2.8 | 0.4 | $1.9 \cdot 10^2$ | $2.9 \cdot 10^2$ | $4.5 \cdot 10^2$ | 2.6 | 7.1 | 16 |
| | 334 | S | AFGINRY | 1.3 | 0.44 | $1 \cdot 10^2$ | $1.7 \cdot 10^2$ | $2.5 \cdot 10^2$ | 1.4 | 4 | 9.5 |
| 10E8 | 671 | KNS | RT | 0.41 | 0.51 | 38 | $1.1 \cdot 10^2$ | $2.2 \cdot 10^2$ | 0.68 | 2.5 | 7.3 |
| | 673 | F | LV | 1.4 | 0.47 | $1.5 \cdot 10^2$ | $2.9 \cdot 10^2$ | $5.1 \cdot 10^2$ | 2.4 | 7 | 18 |
| 3BNC117 | 279 | DN | AHK | 0.33 | 0.22 | $1.9 \cdot 10^2$ | $4.7 \cdot 10^2$ | $9.3 \cdot 10^2$ | 3.4 | 11 | 31 |
| | 281 | AP | TV | 1.1 | 1.1 | 62 | $1.4 \cdot 10^2$ | $2.5 \cdot 10^2$ | 0.97 | 3.2 | 8.6 |
| | 282 | KY | ENR | 1.2 | 0.8 | $1.2 \cdot 10^2$ | $2.3 \cdot 10^2$ | $4 \cdot 10^2$ | 1.9 | 5.6 | 14 |
| | 459 | G | DN | 0.5 | 1 | 54 | $1.4 \cdot 10^2$ | $2.7 \cdot 10^2$ | 0.91 | 3.3 | 9.5 |
| PG9 | 160 | N | KY | 0.4 | 0.2 | $1.5 \cdot 10^2$ | $2.8 \cdot 10^2$ | $4.8 \cdot 10^2$ | 2.3 | 6.7 | 17 |
| | 162 | ST | AIP | 1.2 | 1.1 | $5.4 \cdot 10^2$ | $1.6 \cdot 10^3$ | $2.6 \cdot 10^3$ | 9.5 | 34 | 98 |
| | 169 | GIKMRVW | ELT | 0.53 | 0.92 | $2.4 \cdot 10^2$ | $5.9 \cdot 10^2$ | $1.2 \cdot 10^3$ | 4.1 | 14 | 41 |
| | 171 | KPR | AEGHNQST | 1.4 | 0.62 | $1 \cdot 10^2$ | $1.9 \cdot 10^2$ | $3.1 \cdot 10^2$ | 1.5 | 4.5 | 11 |
| PGT121 | 330 | FHLRSY | Q | 0.12 | 1.4 | 7.4 | $1.6 \cdot 10^2$ | $5.3 \cdot 10^2$ | 0.15 | 3.6 | 16 |
| | 332 | AENV | DIKT | 1.1 | 1.2 | 80 | $1.8 \cdot 10^2$ | $3.2 \cdot 10^2$ | 1.3 | 4.2 | 11 |
| | 334 | DS | GN | 1 | 2 | 15 | 80 | $1.7 \cdot 10^2$ | 0.26 | 1.8 | 5.9 |
| PGT145 | 121 | K | E | 1 | 1 | 34 | 92 | $1.7 \cdot 10^2$ | 0.58 | 2.1 | 5.8 |
| | 160 | N | KY | 0.4 | 0.2 | $1.4 \cdot 10^2$ | $2.5 \cdot 10^2$ | $4.4 \cdot 10^2$ | 2.1 | 6.2 | 15 |
| | 162 | ST | AIP | 1.2 | 1.1 | $6.8 \cdot 10^2$ | $1.2 \cdot 10^3$ | $2 \cdot 10^3$ | 11 | 29 | 73 |
| | 166 | KR | AEGST | 0.73 | 0.58 | 58 | $1.2 \cdot 10^2$ | $2 \cdot 10^2$ | 0.9 | 2.8 | 7.2 |
| | 169 | GIKLTV | EMRW | 0.59 | 1.4 | 8.8 | 80 | $1.9 \cdot 10^2$ | 0.16 | 1.8 | 6.1 |
| PGT151 | 512 | A | GT | 1.1 | 0.57 | $8.6 \cdot 10^2$ | $2.7 \cdot 10^3$ | $4.1 \cdot 10^3$ | 16 | 59 | $1.6 \cdot 10^2$ |
| | 611 | GN | DS | 1.3 | 2 | $3.6 \cdot 10^2$ | $8.9 \cdot 10^2$ | $1.3 \cdot 10^3$ | 6.5 | 20 | 50 |
| | 613 | ST | CN | 0.28 | 0.7 | $1.2 \cdot 10^3$ | $2.8 \cdot 10^3$ | $3.5 \cdot 10^3$ | 21 | 58 | $1.4 \cdot 10^2$ |
| | 637 | DN | EKST | 0.83 | 0.41 | $1.2 \cdot 10^2$ | $2.1 \cdot 10^2$ | $3.5 \cdot 10^2$ | 1.8 | 5.1 | 13 |
| | 639 | T | IM | 1 | 1 | $1.1 \cdot 10^3$ | $1.6 \cdot 10^3$ | $2.6 \cdot 10^3$ | 16 | 42 | 96 |
| VRC01 | 197 | DN | S | 0.5 | 1 | $7.3 \cdot 10^2$ | $1.6 \cdot 10^3$ | $2.3 \cdot 10^3$ | 12 | 35 | 88 |
| | 279 | DN | AHK | 0.33 | 0.22 | $2 \cdot 10^2$ | $4.2 \cdot 10^2$ | $9.4 \cdot 10^2$ | 3.2 | 10 | 30 |
| | 280 | N | D | 1 | 1 | $4.4 \cdot 10^2$ | $1 \cdot 10^3$ | $1.8 \cdot 10^3$ | 7.3 | 24 | 63 |
| | 281 | AP | TV | 1.1 | 1.1 | 52 | $1.3 \cdot 10^2$ | $2.5 \cdot 10^2$ | 0.9 | 3.1 | 8.6 |
| | 458 | G | D | 0.5 | 1 | $2.9 \cdot 10^2$ | $1.2 \cdot 10^3$ | $2.2 \cdot 10^3$ | 6 | 26 | 75 |
| VRC34 | 512 | A | GT | 1.1 | 0.57 | $7.2 \cdot 10^2$ | $1.4 \cdot 10^3$ | $2.9 \cdot 10^3$ | 11 | 34 | 92 |
| | 524 | GP | AERS | 1.5 | 0.67 | 45 | 95 | $1.5 \cdot 10^2$ | 0.71 | 2.2 | 5.6 |
| | 88 | N | K | 0.26 | 0.26 | $5.7 \cdot 10^2$ | $1.4 \cdot 10^3$ | $1.9 \cdot 10^3$ | 9.8 | 30 | 74 |
| | 90 | ST | AEK | 0.48 | 0.8 | $1.4 \cdot 10^3$ | $2.5 \cdot 10^3$ | $3.3 \cdot 10^3$ | 19 | 57 | $1.3 \cdot 10^2$ |

## References

[1] Walker LM, et al. (2009) Broad and potent neutralizing an-tibodies from an African donor reveal a new HIV-1 vaccine target. Science 326: 285–289.

[2] Walker LM, et al. (2011) Broad neutralization coverage of HIV by multiple highly potent antibodies. Nature 477: 466–470.

[3] Liao HX, et al. (2013) Co-evolution of a broadly neutralizing HIV-1 antibody and founder virus. Nature 496: 469–476.

[4] Mouquet H, Nussenzweig MC (2013) HIV: Roadmaps to a vaccine. Nature 496: 441–442.

[5] Klein F, et al. (2013) Antibodies in HIV-1 vaccine development and therapy. Science 341: 1199–1204.

[6] Kwong PD, Mascola JR, Nabel GJ (2013) Broadly neutralizing antibodies and the search for an HIV-1 vaccine: the end of the beginning. Nature Rev Immunol 13: 693–701.

[7] Caskey M, et al. (2015) Viraemia suppressed in HIV-1-infected humans by broadly neutralizing antibody 3BNC117. Nature 522: 487–491.

[8] Caskey M, et al. (2017) Antibody 10-1074 suppresses viremia in HIV-1-infected individuals. Nat Med 23: 185–191.

[9] Bar-On Y, et al. (2018) Safety and anti-viral activity of combination HIV-1 broadly neutralizing antibodies in viremic individuals. Nature medicine 24: 1701–1707.

[10] Sok D, Burton DR (2018) Recent progress in broadly neutralizing antibodies to HIV. Nature immunology 19: 1179–1188.

[11] Zwick MB, et al. (2001) Broadly neutralizing antibodies targeted to the membrane-proximal external region of human immunodeficiency virus type 1 glycoprotein gp41. Journal of virology 75: 10892–10905.

[12] Burton DR, Poignard P, Stanfield RL, Wilson IA (2012) Broadly neutralizing antibodies present new prospects to counter highly antigenically diverse viruses. Science 337: 183–186.

[13] Sparrow E, Friede M, Sheikh M, Torvaldsen S, Newall AT (2016) Passive immunization for influenza through antibody therapies, a review of the pipeline, challenges and potential applications. Vaccine 34: 5442–5448.

[14] Ekiert DC, Wilson IA (2012) Broadly neutralizing antibodies against influenza virus and prospects for universal therapies. Curr Opin Virol 2: 134–141.

[15] Durham ND, et al. (2019) Broadly neutralizing human anti-bodies against dengue virus identified by single B cell tran-scriptomics. eLife 8: 1113.

[16] Chen L, et al. (2009) Structural basis of immune evasion at the site of CD4 attachment on HIV-1 gp120. Science 326: 1123–1127.

[17] Zhou T, et al. (2010) Structural basis for broad and potent neutralization of HIV-1 by antibody VRC01. Science 329: 811–817.

[18] West AP, et al. (2014) Structural insights on the role of anti-bodies in HIV-1 vaccine and therapy. Cell 156: 633–648.

[19] Burton DR, Hangartner L (2016) Broadly Neutralizing An-tibodies to HIV and their role in vaccine design. Annu Rev Immunol 34: 635–659.

[20] Caskey M, Klein F, Nussenzweig MC (2016) Broadly Neutralizing Antibodies for HIV-1 Prevention or Immunotherapy. N Engl J Med 375: 2019–2021.

[21] Gruell H, Klein F (2018) Antibody-mediated prevention and treatment of HIV-1 infection. Retrovirology 15: 73.

[22] Horwitz JA, et al. (2013) HIV-1 suppression and durable control by combining single broadly neutralizing antibodies and antiretroviral drugs in humanized mice. Proc Natl Acad Sci USA 110: 16538–16543.

[23] Shingai M, et al. (2013) Antibody-mediated immunotherapy of macaques chronically infected with SHIV suppresses viraemia. Nature 503: 277–280.

[24] Barouch DH, et al. (2013) Therapeutic efficacy of potent neutralizing HIV-1-specific monoclonal antibodies in SHIV-infected rhesus monkeys. Nature 503: 224–228.

[25] Julg B, et al. (2017) Virological Control by the CD4-Binding Site Antibody N6 in Simian-Human Immunodeficiency Virus-Infected Rhesus Monkeys. J Virol 91: 1633.

[26] Bar KJ, et al. (2016) Effect of HIV Antibody VRC01 on Viral Rebound after Treatment Interruption. N Engl J Med 375: 2037–2050.

[27] Lienhardt C, et al. (2012) New drugs for the treatment of tuberculosis: needs, challenges, promise, and prospects for the future. J Infect Dis 205: S241–9.

[28] Yu WH, et al. Predicting the broadly neutralizing antibody susceptibility of the HIV reservoir. JCI Insight 4: e130153.

[29] Wagh K, et al. (2016) Optimal Combinations of Broadly Neu-tralizing Antibodies for Prevention and Treatment of HIV-1 Clade C Infection. PLOS Pathogens 12: e1005520.

[30] Lu CL, et al. (2016) Enhanced clearance of HIV-1-infected cells by broadly neutralizing antibodies against HIV-1 in vivo. Science 352: 1001–1004.

[31] Reeves DB, et al. (2020) Mathematical modeling to reveal breakthrough mechanisms in the HIV Antibody Mediated Prevention (AMP) trials. PLoS Comput Biol 16: e1007626.

[32] Saha A, Dixit NM (2020) Pre-existing resistance in the latent reservoir can compromise VRC01 therapy during chronic HIV-1 infection. PLoS Comput Biol 16: e1008434.

[33] Meijers M, Vanshylla K, Gruell H, Klein F, Lässig M (2021) Predicting in vivo escape dynamics of HIV-1 from a broadly neutralizing antibody. Proceedings of the National Academy of Sciences 118: e2104651118.

[34] Lemey P, Rambaut A, Pybus OG (2006) HIV evolutionary dynamics within and among hosts. AIDS Rev 8: 125–140.

[35] Zanini F, et al. (2015) Population genomics of intrapatient HIV-1 evolution. eLife 4: e11282.

[36] Feder AF, et al. (2016) More effective drugs lead to harder selective sweeps in the evolution of drug resistance in HIV-1. eLife 5: 1161.

[37] Nourmohammad A, Otwinowski J, Luksza M, Mora T, Wal-czak AM (2019) Fierce Selection and Interference in B-Cell Repertoire Response to Chronic HIV-1. Mol Biol Evol 36: 2184–2194.

[38] Neher RA, Leitner T (2010) Recombination rate and selection strength in HIV intra-patient evolution. PLoS Comput Biol 6: e1000660.

[39] Bonhoeffer S, Chappey C, Parkin NT, Whitcomb JM, Petropoulos CJ (2004) Evidence for positive epistasis in HIV-1. Science 306: 1547–1550.

[40] Zhang TH, et al. (2020) Predominance of positive epistasis among drug resistance-associated mutations in HIV-1 pro-tease. PLoS Genet 16: e1009009.

[41] Ferguson AL, et al. (2013) Translating HIV sequences into quantitative fitness landscapes predicts viral vulnerabilities for rational immunogen design. Immunity 38: 606–617.

[42] Dingens AS, Arenz D, Weight H, Overbaugh J, Bloom JD (2019) An Antigenic Atlas of HIV-1 Escape from Broadly Neu-tralizing Antibodies Distinguishes Functional and Structural Epitopes. Immunity 50: 520–532.e3.

[43] Perelson AS, Neumann AU, Markowitz M, Leonard JM, Ho DD (1996) HIV-1 dynamics in vivo: virion clearance rate, in-fected cell life-span, and viral generation time. Science 271: 1582–1586.

[44] Lynch RM, et al. (2015) HIV-1 Fitness Cost Associated with Escape from the VRC01 Class of CD4 Binding Site Neutralizing Antibodies. Journal of Virology 89: 4201–4213.

[45] Scheid JF, et al. (2016) HIV-1 antibody 3BNC117 suppresses viral rebound in humans during treatment interruption. Nature 535: 556–560.

[46] Pancera M, Changela A, Kwong PD (2017) How HIV-1 entry mechanism and broadly neutralizing antibodies guide structure-based vaccine design. Curr Opin HIV AIDS 12: 229–240.

[47] Dingens AS, Haddox HK, Overbaugh J, Bloom JD (2017) Comprehensive Mapping of HIV-1 Escape from a Broadly Neutralizing Antibody. Cell Host Microbe 21: 777–787.e4.

[48] Schommers P, et al. (2020) Restriction of HIV-1 Escape by a Highly Broad and Potent Neutralizing Antibody. Cell 180: 471–489.e22.

[49] Zanini F, Puller V, Brodin J, Albert J, Neher RA (2017) In vivo mutation rates and the landscape of fitness costs of HIV-1. Virus Evol 3: vex003.

[50] Perelson AS (2002) Modelling viral and immune system dynamics. Nature Rev Immunol 2: 28–36.

[51] Meintjes PL, Rodrigo AG (2005) Evolution of relative synonymous codon usage in Human Immunodeficiency Virus type-1. J Bioinform Comput Biol 3: 157–168.

[52] Gruell H, Klein F (2017) Progress in HIV-1 antibody research using humanized mice. Curr Opin HIV AIDS 12: 285–293.

## References

[1] Caskey M, et al. (2015) Viraemia suppressed in HIV-1-infected humans by broadly neutralizing antibody 3BNC117. Nature 522: 487–491.

[2] Caskey M, et al. (2017) Antibody 10-1074 suppresses viremia in HIV-1-infected individuals. Nat Med 23: 185–191.

[3] Bar-On Y, et al. (2018) Safety and anti-viral activity of combination HIV-1 broadly neutralizing antibodies in viremic individuals. Nature medicine 24: 1701–1707.

[4] Zanini F, et al. (2015) Population genomics of intrapatient HIV-1 evolution. eLife 4: e11282.

[5] Gaschen B, Kuiken C, Korber B, Foley B (2001) Retrieval and on-the-fly alignment of sequence fragments from the HIV database. Bioinformatics (Oxford, England) 17: 415–418.

[6] Dingens AS, Arenz D, Weight H, Overbaugh J, Bloom JD (2019) An Antigenic Atlas of HIV-1 Escape from Broadly Neutralizing Antibodies Distinguishes Functional and Structural Epitopes. Immunity 50: 520–532.e3.

[7] Scheid JF, et al. (2016) HIV-1 antibody 3BNC117 suppresses viral rebound in humans during treatment interruption. Nature 535: 556–560.

[8] Zhou T, et al. (2015) Structural repertoire of HIV-1-neutralizing antibodies targeting the CD4 supersite in 14 donors. Cell 161: 1280–1292.

[9] LaBranche CC, et al. (2018) HIV-1 envelope glycan modifications that permit neutralization by germline-reverted VRC01-class broadly neutralizing antibodies. PLoS pathogens 14: e1007431.

[10] Lynch RM, et al. (2015) HIV-1 Fitness Cost Associated with Escape from the VRC01 Class of CD4 Binding Site Neutralizing Antibodies. Journal of Virology 89: 4201–4213.

[11] Horwitz JA, et al. (2013) HIV-1 suppression and durable control by combining single broadly neutralizing antibodies and antiretroviral drugs in humanized mice. Proceedings of the National Academy of Sciences 110: 16538–16543.

[12] Otsuka Y, et al. (2018) Diverse pathways of escape from all well-characterized VRC01-class broadly neutralizing HIV-1 antibodies. PLOS Pathogens 14: e1007238.

[13] García F, et al. (1999) Dynamics of viral load rebound and immunological changes after stopping effective antiretroviral therapy. Aids 13: F79–F86.

[14] Ioannidis JP, et al. (2000) Dynamics of HIV-1 viral load rebound among patients with previous suppression of viral replication. Aids 14: 1481–1488.

[15] McCullagh P, Nelder JA (2019) Generalized Linear Models. Boca Raton: Routledge.

[16] Wilkinson DJ (2019) Stochastic Modelling for Systems Biology. Chapman & Hall/CRC Mathematical and Computational Biology. Boca Raton: CRC Press, Taylor & Francis Group, third edition.

[17] Gillespie DT (1977) Exact stochastic simulation of coupled chemical reactions. The journal of physical chemistry 81: 2340–2361.

[18] Risken H (1996) The Fokker-Planck Equation: Methods of Solution and Applications. Number v. 18 in Springer Series in Synergetics. New York: Springer-Verlag, second edition.

[19] Crow JF, Kimura M (2010) An Introduction to Population Genetics Theory. Caldwell: The Blackburn Press.

[20] Allen LJ (2010) An Introduction to Stochastic Processes with Applications to Biology. Boca Raton: CRC press.

[21] Geman S, Geman D (1984) Stochastic relaxation, Gibbs distributions, and the Bayesian restoration of images. IEEE Transactions on pattern analysis and machine intelligence : 721–741.

[22] García F, et al. (2001) The virological and immunological consequences of structured treatment interruptions in chronic HIV-1 infection. Aids 15: F29–F40.

[23] Zanini F, Puller V, Brodin J, Albert J, Neher RA (2017) In vivo mutation rates and the landscape of fitness costs of HIV-1. Virus Evol 3: vex003.

[24] Nielsen R (2006) Statistical Methods in Molecular Evolution. New York: Springer.

[25] Theys K, et al. (2018) Within-patient mutation frequencies reveal fitness costs of CpG dinucleotides and drastic amino acid changes in HIV. PLoS genetics 14: e1007420.

[26] Feder AF, et al. (2017) A spatio-temporal assessment of simian/human immunodeficiency virus (SHIV) evolution reveals a highly dynamic process within the host. PLoS pathogens 13: e1006358.

[27] Stoddart JA, Taylor JF (1988) Genotypic diversity: Estimation and prediction in samples. Genetics 118: 705–711.

[28] Kimura M (1981) Estimation of evolutionary distances between homologous nucleotide sequences. Proceedings of the National Academy of Sciences 78: 454–458.

[29] Owen A, Zhou Y (2000) Safe and effective importance sampling. Journal of the American Statistical Association 95: 135–143.

[30] Hastings WK (1970) Monte Carlo sampling methods using Markov chains and their applications. Biometrika 57: 97–109.

[31] Chaillon A, et al. (2020) HIV persists throughout deep tissues with repopulation from multiple anatomical sources. The Journal of clinical investigation 130: 1699–1712.

[32] Wong JK, Yukl SA (2016) Tissue Reservoirs of HIV. Current opinion in HIV and AIDS 11: 362–370.

[33] Chun TW, et al. (2005) HIV-infected individuals receiving effective antiviral therapy for extended periods of time continually replenish their viral reservoir. The Journal of clinical investigation 115: 3250–3255.

[34] Avettand-Fenoel V, et al. (2007) Failure of bone marrow transplantation to eradicate HIV reservoir despite efficient HAART. Aids 21: 776–777.

[35] Shan L, Siliciano RF (2013) From reactivation of latent HIV-1 to elimination of the latent reservoir: The presence of multiple barriers to viral eradication. Bioessays 35: 544–552.

[36] Sharkey M, et al. (2011) Episomal viral cDNAs identify a reservoir that fuels viral rebound after treatment interruption and that contributes to treatment failure. PLoS pathogens 7: e1001303.

[37] Tagarro A, et al. (2018) Early and highly suppressive ART are main factors associated with low viral reservoir in european perinatally HIV infected children. Journal of acquired immune deficiency syndromes (1999) 79: 269.

[38] Ekström M (2008) Alternatives to maximum likelihood estimation based on spacings and the Kullback–Leibler divergence. Journal of Statistical Planning and Inference 138: 1778–1791.

[39] Lindsay BG (1994) Efficiency versus robustness: The case for minimum Hellinger distance and related methods. The annals of statistics 22: 1081–1114.

[40] Simpson DG (1987) Minimum Hellinger distance estimation for the analysis of count data. Journal of the American statistical Association 82: 802–807.

[41] Graham RL, Knuth DE, Patashnik O, Liu S (1989) Concrete mathematics: A foundation for computer science. Computers in Physics 3: 106–107.

[42] Feder AF, Pennings PS, Petrov DA (2021) The clarifying role of time series data in the population genetics of HIV. PLoS genetics 17: e1009050.

[43] Nourmohammad A, Otwinowski J, Luksza M, Mora T, Walczak AM (2019) Fierce Selection and Interference in B-Cell Repertoire Response to Chronic HIV-1. Mol Biol Evol 36: 2184–2194.

[44] Feder AF, et al. (2016) More effective drugs lead to harder selective sweeps in the evolution of drug resistance in HIV-1. eLife 5: 1161.

[45] Strelkowa N, Lässig M (2012) Clonal interference in the evolution of influenza. Genetics 192: 671–682.

[46] Hill WG, Robertson A (1966) The effect of linkage on limits to artificial selection. Genetics Research 8: 269–294.

[47] Comeron JM, Williford A, Kliman RM (2008) The Hill–Robertson effect: Evolutionary consequences of weak selection and linkage in finite populations. Heredity 100: 19–31.

[48] Fisher RA (1956) Statistical Methods and Scientific Inference. London: Oliver and Boyd, first edition.

[49] Fisher RA (1954) Statistical Methods for Research Workers. New York: Hafner Publishing, twelfth edition.

[50] Spira S, Wainberg MA, Loemba H, Turner D, Brenner BG (2003) Impact of clade diversity on HIV-1 virulence, antiretroviral drug sensitivity and drug resistance. Journal of Antimicrobial Chemotherapy 51: 229–240.

